# Development and Validation of a Paralimbic Related Subcortical Brain Dysmaturation MRI Score in Infants with Congenital Heart Disease

**DOI:** 10.1101/2024.04.21.24306144

**Authors:** William T. Reynolds, Jodie K. Votava-Smith, George Gabriel, Vince Lee, Vidya Rajagopalan, Yijen Wu, XiaoQin Liu, Hisato Yagi, Ruby Slabicki, Brian Gibbs, Nhu N. Tran, Molly Weisert, Laura Cabral, Subramanian Subramanian, Julia Wallace, Sylvia del Castillo, Tracy Baust, Jacqueline Weinberg, Lauren Lorenzi Quigley, Jenna Gaesser, Sharon H. O’Neil, Vanessa Schmithorst, Rafael Ceschin, Cecilia Lo, Ashok Panigrahy

## Abstract

**Background:** Brain magnetic resonance imaging (MRI) of infants with congenital heart disease (CHD) shows brain immaturity assessed via a cortical-based semi-quantitative score. Our primary aim was to develop an infant paralimbic-related subcortical-based semi-quantitative dysmaturation score, a brain dysplasia score (BDS), to detect abnormalities in CHD infants and predict clinical outcomes. Our secondary aim was to validate our BDS in a preclinical mouse model of hypoplastic left heart syndrome.

**Methods:** A paralimbic-related subcortical BDS, derived from structural MRIs of infants with CHD, was correlated with clinical risk factors, regional cerebral volumes, feeding and 18-month neurodevelopmental outcomes. The BDS was validated in a known CHD mouse model named *Ohia* with two disease-causing genes, *Sap130* and *Pchda9*. To relate clinical findings, RNA-Seq was completed on *Ohia* animals.

**Findings:** BDS showed high incidence of paralimbic-related subcortical abnormalities (including olfactory, cerebellar, and hippocampal abnormalities) in CHD infants (n=215) compared to healthy controls (n=92). BDS correlated with reduced cortical maturation, developmental delay, poor language and feeding outcomes, and increased length of stay. *Ohia* animals (n=63) showed similar BDS findings, and RNA-Seq analysis showed altered neurodevelopmental and feeding pathways. *Sap130* mutants correlated with a more severe BDS whereas *Pcdha9* correlated with a milder phenotype.

**Interpretation:** Our BDS is sensitive to dysmaturational differences between CHD and healthy controls, and predictive of poor outcomes. A similar spectrum of paralimbic-related subcortical abnormalities exists between human and *Ohia* mutants suggesting a common genetic mechanistic etiology.

**Funding:** National Library of Medicine, Department of Defense, National Heart, Lung, and Blood Institute, National Institute on Aging, Southern California Clinical and Translational Sciences Institute, Additional Ventures Foundation, Saban Research Institute, Children’s Hospital Los Angeles Clinical Services Research Grant, and National Institute of Nursing Research. Funding award numbers can be found in the acknowledgment section.

**Research In Context:** *Evidence before:* The number of clinical and research MRI studies in neonatal/infant CHD subjects has increased dramatically in the last two decades. Previous studies have developed brain MRI scores that have focused on cortical structural maturation and acquired brain injury. Paralimbic-related subcortical regions are important for the development of cognitive and visuomotor functions in early development. Levering a large infant brain MRI dataset and a large-scale genetic mouse screen, we theorized that a paralimbic-related subcortical brain MRI score could assist clinicians with outcome prediction in CHD infants.

*Added Value:* This work aims to develop a subcortical morphological scoring system that could be applied to either clinical or research MRI scans and could improve the ability of clinicians and neuroradiologists to predict not only those at risk for suboptimal neurodevelopmental outcomes but also associated co-morbidities. We discovered not only are there paralimbic-related subcortical structural abnormalities that a brain MRI score can detect but also that this score predicted poor language outcomes, poor feeding outcomes, and increased post-surgical length of stay. We also found that the genetic model of hypoplastic left heart syndrome, the most severe form of CHD, also demonstrated a similar pattern of paralimbic related subcortical brain abnormalities.

*Implications:* This novel scoring system developed by our group has implications for early detection of at-risk CHD individuals for poor outcomes, both neurodevelopmental and quality of life. This subcortical paralimbic brain dysplasia score is a simple tool that can be easily added to neuroradiological workflows that can lead to better outcome prediction for children with CHD. Our scoring system helps us to better serve our population, allowing clinicians and researchers to prognosticate highest risk individuals who will benefit from the earliest forms of intervention.

## Introduction

Congenital heart disease (CHD) affects 1% of live births each year.^1,2^ As the surgical care for CHD improves, there remains lingering increased risk of poor neurodevelopmental outcomes for patients with CHD across the lifespan. The mechanism for neurodevelopmental disabilities in non-syndromic CHD is unknown and is thought to be related to fetal exposure to reduced substrate delivery/hypoxia or a genetic underpinning. Genetic abnormalities are detected in approximately 50% of children with syndromic CHD, and in approximately 10% of children without a recognizable clinical phenotype.^3^ Significant overlap is present between deleterious de novo mutations and previously reported mutations associated with neurodevelopmental disorders,^4^ suggesting that genetic mutations that cause CHD, may also be important in the etiology of neurodevelopmental deficits.^5,6^ Smaller brain volumes and dysmature brain structures are seen in neonates and fetuses with CHD before cardiac surgery, suggesting that innate and/or prenatal factors may play an important role in altering brain development.^7–9^ Within our current understanding of neurogenesis, there is little known about the role that deep grey and subcortical regions play in mediating poor neurodevelopmental outcomes in relation to genetic alterations. Recent animal models and correlative neuropathological studies suggest that cortical dysmaturation is linked to white matter abnormalities, including developmental vulnerability of the subplate in CHD.^10,11^ Recent neuroimaging studies have documented paralimbic related subcortical morphological abnormalities in CHD patients across the lifespan,^12–16^ but the relationship between paralimbic-related subcortical morphological abnormalities and clinical and neurodevelopmental outcomes in CHD is also unknown.^17–20^

Children with CHD are at a higher risk of developing brain dysmaturation, a generalized term encompassing abnormal and delayed development of brain macro- and microstructure^21–25^. CHD patients are also at risk for acquired brain injury, including infarcts and small vessel disease across lifespan as detected by conventional neuroimaging studies. Clinical neuroimaging studies are also becoming more common to obtain in patients during the peri-operative period based on recently published guidelines. Most conventional semi-quantitative MRI scoring systems in CHD patients have exclusively focused on either acquired brain injury or cortical maturation assessment, particularly in the neonatal period.^26–33^ We recently developed and validated an infant semi-quantitative score that extending beyond cortical maturation and acquired brain injury to include morphological alterations in paralimbic-related subcortical structures (including cerebellum, hippocampus, olfactory bulb abnormalities ), CSF-related abnormalities (including increased extra-axial CSF in frontotemporal regions) and the corpus callosum known as a brain dysplasia score (BDS), informed by preclinical models of CHD, specifically hypoplastic left heart syndrome (HLHS). We focused our analysis on paralimbic-related subcortical structures that are known to undergo neurogenesis in early development and across the lifespan, including the olfactory bulbs, hippocampus, and cerebellum. We have recently described a similar pattern of structural subcortical dysmaturation both in human infants with CHD and genetically relevant ciliary motion dysfunction, and also in relation to preclinical models of CHD including hypoplastic left heart syndrome (HLHS).^34–40^ We have previously shown that the BDS correlates with abnormal neonatal brain white matter connectivity patterns^41^ but have yet to validate this scoring system in a large dataset in relation to clinical and neurodevelopmental outcomes. Here, we used quantitative structural brain magnetic resonance imaging (MRI) in infants with CHD to test the hypothesis that subcortical morphological measurements could be assessed using a qualitative scoring system termed brain dysplasia score (BDS) and that these subcortical structures are potential predictors of not only infant cortical maturation and regional brain volumes but also are predictors of poor clinical and neurodevelopmental outcomes.^12–15,21,42,43^

To further validate our subcortical BDS from a genetic perspective, we leveraged the *Ohia* mouse model of HLHS recovered from a large-scale mouse mutagenesis screen. In *Ohia* mice with HLHS, phenotypic mutations primarily arise from homozygous mutations in two genes, a chromatin modifying protein Sin3A-associated protein 130 (*Sap130*), and protocadherin A9 (*Pcdha9*), a cell adhesion protein in the a-protocadherin gene cluster.^44^ Importantly, both genes are known to have important roles in brain health-related outcomes. For example, the clustered protocadherins provide cell surface diversity by encoding unique neuronal identity essential for patterning synaptic connectivity.^45^ Mice with PCDHA mutations have deficiencies in both brain connectivity and neurobehavioral deficits.^46^ Moreover, mutations in both PCDHA and SIN3A are associated with autism and Rett Syndrome in humans.^47–49^ Importantly, the SIN3A complex also contains transcription factors known to regulate developmental and lifespan neurogenesis.^50,51^ Given the prominent roles of PCDHA and SIN3A in human neurogenesis and brain development, we used the *Ohia* mouse model for further validation of the hypothesis that the subcortical BDS could be used as a proxy of subcortical morphological and connectivity quantitative measurements and predict clinical and neurodevelopmental outcomes in human CHD infants.

## Methods

### Ethics

The study was HIPAA compliant and approved by the institutional review board (IRB) of Children’s Hospital Los Angeles (CHLA) and Children’s Hospital of Pittsburgh (CHP) of the University of Pittsburgh Medical Center (UPMC) and written informed consent was obtained from each subject, except for a few patients in which MRI was obtained clinically, and retrospective use of the data was approved by the IRB protocol (CHLA-CCI-09-00055, CCI-10-00235, University of Pittsburgh-20030213,19100215, 19030204). All experiments were performed per the institutional guidelines and regulations.

### Patient Recruitment and Clinical Data Collection

This study is a secondary analysis of neonates with CHD (term and preterm) undergoing brain MRI scans who were recruited as part of a prospective, observational study from two large medical centers, CHLA and CHP. Neonatal CHD cases were prospectively recruited from (1) pregnancies with fetal CHD confirmed with fetal echocardiography and (2) postnatal admissions to the cardiothoracic intensive care unit postoperatively between 2002-2016 and (3) infants with both preterm and term CHD undergoing clinically indicated brain MRI scans at approximately term-equivalent gestational age (GA) were recruited both prospectively and retrospectively in the peri-operative period from 2002-2017. The inclusion criterion was critical CHD defined as a heart defect expected to require corrective or palliative cardiac surgery during infancy. Exclusion criteria for the CHD included documented major chromosomal abnormalities and major congenital brain malformations. Normal referents were recruited (1) from healthy pregnant volunteers and (2) postnatally from a normal newborn nursery. An additional comparison group was included which was comprised of preterm infants without CHD who were recruited from a high-risk NICU at the same institutions as previously published.^20,52–54^ Details about the recruitment strategies have been described in multiple previous publications.^12,34,35,55–63^

There were 423 subjects that met the inclusion criteria for this study: 307 full term neonates (215 with CHD and 92 term controls) and 116 preterm neonates (65 with CHD born 25-36 weeks GA, and 51 preterm control patients born 24 – 36 weeks GA) (Figure 1). This is the dataset for which we performed the primary analysis of calculating a BDS and comparing the difference between groups.

**Figure 1.**
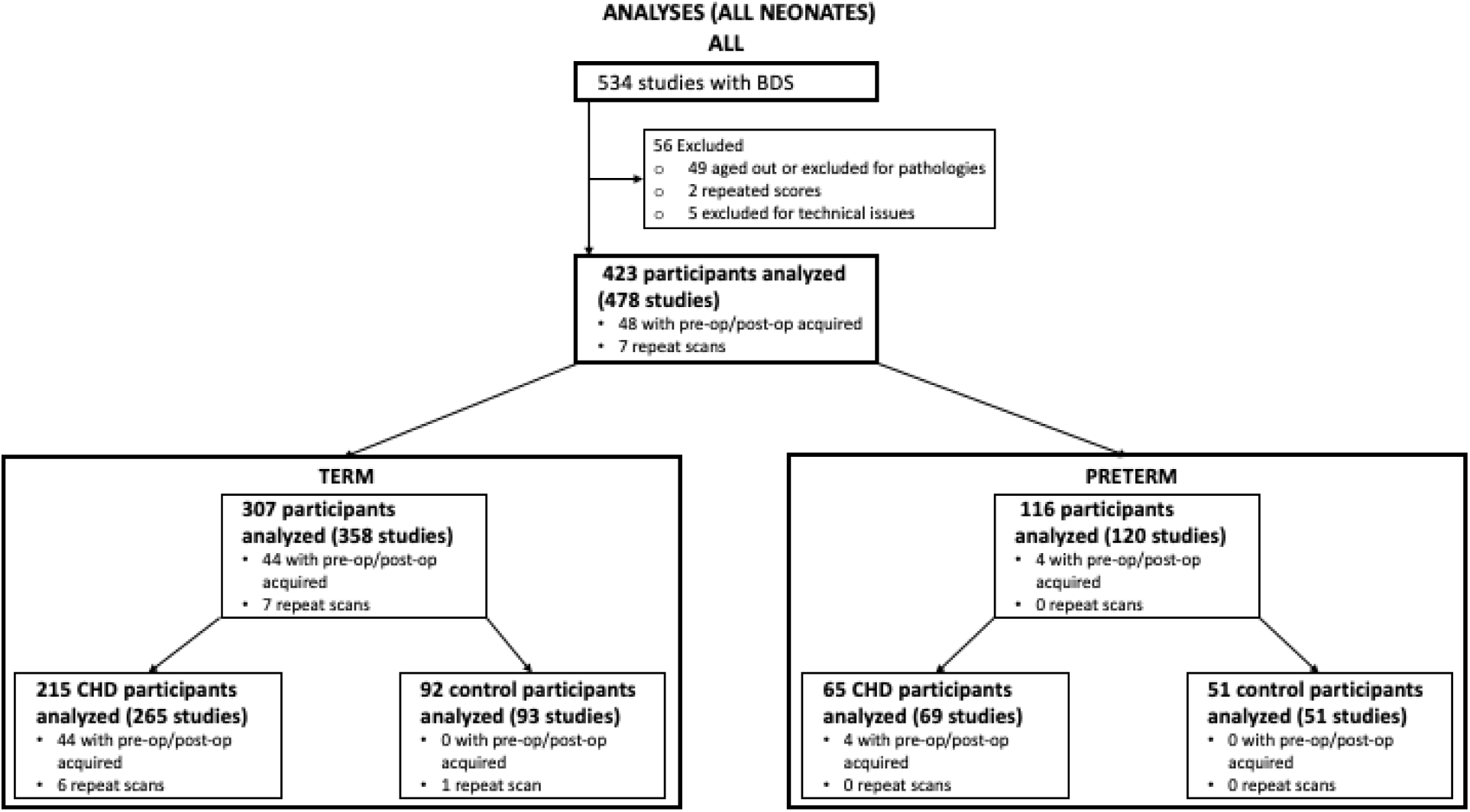
Recruitment Flowchart and Subject Numbers. Study initially consisted of 534 subjects which were scored using the brain dysplasia score (BDS). Initial subjects consisted of 307 term and 116 pre-term individuals.

### Research Scans

Brain MRIs for CHD infants were obtained in either the preoperative (usually between 1-7 days) or postoperative period (up to 13 weeks postnatally, but often sooner). The age range for healthy controls ranged between birth and thirteen weeks postnatally, encompassing the same range of the CHD infants. Preoperative research brain imaging was conducted when on CHD subjects when the cardiothoracic intensive care unit (CTICU) team/cardiology team determined the patient was stable for transport to the MRI scanner. Postoperative research scan was performed at less than three months of postnatal age either as an inpatient or outpatient. Most of our scans were research indicated and as such no additional sedation/anesthesia was given for the scan purpose. Most of the pre-operative scans were performed on non-intubated non-sedated patients; however, if the patient was intubated and sedated for clinical reasons at the time of the scan, their clinically indicated sedation continued under care of the primary CTICU. team. The post-operative scans were performed when the infant was clinically stable and thus were done as “feed and bundle” scans without sedation. To minimize movement during imaging, infants were secured in Med-Vac Immobilization Bag (CFI Medical) with multiple levels of ear protection, including ear plugs, MiniMuffs (Natus Medical Incorporated), and standard headphones.

### Clinical Scans

Patients in the clinically indicated brain MRI group were scanned at approximately term-equivalent GA either in the pre- or post-operative period as previously published.^58^ The post-operative scan for both preterm CHD and term CHD groups was performed at least within 52 weeks corrected postconceptional age.

### Neonatal Brain MRI Protocol

MRI studies were acquired: (1) GE 1.5T (Signa LX, GE Healthcare, Milwaukee, WI) MR system using a custom-built neonatal transmit-receive head coil, (2) Philips 3T Achieva MR System (Ver. 3.2.1.1) using standard 8-channel SENSE head coil; and (3) Siemens Skyra 3T scanner using a 20-channel coil. Conventional imaging studies were acquired with the MRS studies and included a 3D coronal SPGR sequence (TE= 6 ms; TR=25 ms, FOV=18 cm; matrix=256 × 160; slice thickness 1.5 mm, spacing 0 mm) or axial and sagittal T1-weighted FLAIR sequences (TE=7.4, TR=2100; TI=750; FOV=20 cm; Matrix=256 × 160), axial T2-weighted FSE sequence (TE=85ms, TR=5000ms, FOV=20 cm, matrix =320 × 160 or 256 × 128) and a diffusion-weighted sequence (TE=80; TR=10000; FOV=22 cm; Matrix = 128 × 128; slice thickness =4.5 mm, spacing 0 mm). The 3D T1-weighted, T2-weighted, and diffusion-weighted images were reviewed by two pediatric neuroradiologists for evidence of punctate white matter lesion, hypoxic-ischemic injury, acute focal infarction, and hemorrhage as previously described and were used to construct dichotomized and composite brain injury scores.^64^

### Subcortical brain dysplasia MRI score (BDS) derivation

Our development of the subcortical based BDS has been derived from observations from both preclinical mouse CHD mutants and human CHD infants from recent studies from our group.^21,22,36,39,65–67^ The evaluation for brain dysplasia in our human CHD population is conducted by examining the following structures: cerebellar hemispheres (both left and right together), cerebellar vermis, right olfactory bulb, right olfactory sulcus, left olfactory bulb, left olfactory sulcus, hippocampus, choroid plexus, brain stem, corpus callosum, and supratentorial extra-axial fluid. Supplemental Figure 1 depicts a visualization of how the BDS was calculated. Except for the supratentorial extra-axial fluid, each of the structures listed were evaluated on whether they appear normal or abnormal. For cerebellar hemispheres and cerebellar vermis, they were scored for hypoplasia (small volume) and dysplasia (abnormal shape). Likewise, the olfactory bulbs and sulci were evaluated as separate structures for purposes of scoring. Based on the finding, a binary number score is given for each structure, with a score of 0 assigned for normal and a score of 1 for abnormal. For olfactory bulbs and sulci, both hypoplastic and absent findings are treated as abnormal and given a score of 1. For supratentorial extra-axial fluid, the structural features demonstrate different gradations of abnormality – none, mild, moderate, or severe fluid accumulation – and thus successively higher integer scores are assigned as the severity of abnormality becomes greater. The integer scores for each of the structures were then summed to create the BDS with “olfactory correction” which treats both absent and hypoplastic olfactory abnormalities as one score point, thus making the weighting of the olfactory system in line with cerebellum and hippocampus. Higher BDS values can be thought of as having a “worse” score as subjects had more abnormalities present. A subset of cases was reviewed by two raters to validate the scoring.

Similar to our BDS, a brain injury score was calculated for each human subject. The composite brain injury score consisted of four criteria with each criterion being scored as a 1 if present in the subject. The four criteria were hemorrhage, infarct, hypoxic ischemic injury, and periventricular white matter injury (PWMI). A dichotomized version was also calculated for each individual and was scored as a 1 or 0. If the original brain injury score was greater than 0, then the composite score was a 1; otherwise, the score was a 0. A visual depiction of the brain injury scores can be found in Supplemental Figure 2.

### Structural Co-variate Analysis: Cortical Maturation Score and Regional Brain Volumetric Analysis

We correlated BDS with a classic cortical Total Maturation Score (TMS) and regional brain volumes. A cortical TMS was calculated for all subjects with brain MRI included in this analysis. A subset of the full cohort (term controls and CHD scanned at 3T imaging, n=105) underwent cerebral regional volumetric segmentation for structural co-variate analysis. To determine the relationship between our qualitative BDS and quantitative regional morphological dysmaturation, we performed regional brain morphometric techniques including volumetric segmentation of total intracranial cerebral spinal fluid (CSF), cortical grey matter, cortical white matter, deep grey nuclei, brainstem, and cerebellum using a neonatal and infant brain segmentation age-specific atlas. We further segmented total intracranial CSF into three compartments: supratentorial extra-axial CSF, infratentorial extra-axial CSF and intraventricular CSF. Our group used the above age-specific atlas to build a semi-automated brain parcellation pipeline for use in neonates and young infants (Supplemental Figure 3). The automated processing pipeline which was previously described in by our group,^35^ was developed using Nipype and interfaces with several image registration algorithms using age-appropriate neonatal/infant templates to accommodate different post-conceptional ages in the patient cohort. Our methodology for volumetric segmentation of neonatal brain has been previously described.^68,69^ We further refined this methodology for the current study because of the wide range of post-conceptional ages of neuroimaging studies and the resulting need to use a range of neonatal and infant age-appropriate templates. We also performed validation experiments to ensure optimal performance with this neonatal/infant CHD cohort with mild brain dysplasia. We processed each patient’s T1 and T2 volumetric images in parallel to optimize registration parameters uniquely to each tissue contrast. First, the images were cropped, and brain extracted using FSL BET. The volumetric images (T1 and T2 separately) were then biasfield corrected using FSL’s FAST segmentation. We then registered the bias-field corrected images to the tissue contrast specific and post-conceptional age matched template, created by Serag et. al,^70^ using a non-linear registration from Advanced Normalization Tools (ANTs). The output transformations were inversed and applied to the template space tissue probability maps provided with the neonatal atlas. This transformation results in subject-space tissue probability maps. We calculated tissue volumes by thresholding the partial volume maps at a visually acceptable lower bound and extracting the volume from the binarized mask. The subject-space segmentations were manually checked for accuracy by an expert pediatric neuroradiologist (AP) blinded to patient diagnosis.

### Clinical Risk Factor Analysis

We correlated BDS with clinical risk factors. Analysis of clinical risk factors included a subset of subjects with critical CHD who required corrective or palliative cardiac surgery within the first month of life who were prospectively enrolled for pre and postoperative brain MRI scans from 2009-2016. Two hundred ninety-one subjects were prospectively enrolled from June 2009 to October 2016. Of these subjects, 158 met exclusion criteria including 57 with no MRI done, 38 due to prematurity, 38 passed the age threshold, 11 expired preoperatively, 10 had no neonatal surgery and 4 had a postnatal major genetic diagnosis. Of the 133 term CHD infants with brain MRI meeting inclusion criteria, 90 subjects had sufficient imaging quality for structural analysis and clinical risk factor data collected. comprised the study group for this analysis. Subjects that had a known major chromosomal abnormality, were premature (<=36 weeks of age), died prior to MRI or did not require neonatal cardiac surgery were excluded in this analysis. Clinical data was collected from the electronic medical records and included 18 patient-specific and 9 preoperative variables associated with preoperative scan and 6 intra-operative (e.g., cardiopulmonary bypass, deep hypothermic circulatory arrest times) and 12 postoperative variables associated with postoperative scan, as described previously.^59,71^ CHD lesions were classified in several ways (not mutually exclusive) including postnatal cyanosis, presence of aortic arch obstruction, single vs. double ventricles, d-transposition of the great arteries, conotruncal defects, heterotaxy, whether the lesion alters fetal cerebral substrate delivery and severity of this alteration (normal, altered, severely altered).

### Feeding Outcomes

We correlated the BDS with feeding outcomes. The feeding outcome study group consisted of term infants with CHD who had neonatal brain MRI with brain dysplasia score between 2003-2015, enrolled both prospectively for research MRI and retrospectively after clinical MRI, at CHLA and CHP. Subjects with major gastrointestinal anomalies or surgeries, a diagnosis of CHARGE syndrome, death within the first 30 days of life, or death prior to initiation of feeds were excluded from the feeding analysis. For the feeding study, we evaluated 177 term infants with CHD who had MRI that were used to calculate BDS as described above. Of those, 32 were excluded including 14 with major gastrointestinal anomalies or surgeries, 1 with brain anomaly, 6 with CHARGE syndrome, and 11 that died in the first 30 days or prior to initiation of feeds. Therefore, the feeding analysis group consisted of 145 subjects of which 81 were from CHLA and 64 from CHP. The inpatient medical records were assessed for the following feeding-related variables: presence of dysphagia, aspiration, gastroesophageal reflux, gastrointestinal dysmotility, intestinal malrotation, vocal cord paralysis, use of enteral tube feeding (including nasogastric, nasojejunal, or surgically placed gastric or jejunal tubes), lack of oral feeding prior to neonatal hospital discharge, and length of neonatal hospitalization.

### Neurodevelopmental Outcomes

For neurodevelopmental outcomes assessment, Bayley Scales of Infant and Toddler Development or Battelle Developmental Inventory were completed by a licensed psychologist in a subset of patients. A total of 90 subjects (CHLA n=32, CHP n=58) had early neurodevelopmental outcome data obtained between 15 to 18 months postnatal age. The standard score for each developmental domain (motor, language/communication, cognitive, and social/emotional) was evaluated. Developmental delay was defined as a standard score greater than 2 standard deviations below the mean in 1 developmental domain. Global developmental delay was defined as a standard score greater than 2 standard deviations below the mean in 2 or more developmental domains. The following criteria was used: Average= Standard Score 90 to 109 (25^th^ to 74^th^ percentile); Low Average= Standard Score 80 to 89 (9^th^ to 24^th^ percentile); Below Average= (2^nd^ to 8^th^ percentile); Exceptionally low= (<2nd percentile). The data was then dichotomized to yes/no for (1) developmental delay, (2) global developmental delay, and (3) below average for each subject to facilitate harmonization between the two different test exams across both sites.

### Mouse Screen Analysis

A retrospective analysis was carried out on the mouse lines described by Li et al.^72^ In brief, a forward recessive mouse screen was carried out using N-ethyl-N-Nitrosourea in which 3700 mouse lines, defined by G1 sire, were screened for defects. Defect screening was completed through ultrasound scanning, micro-computed tomography (CT), micro-magnetic resonance imaging (MRI) and visual inspection of G3 offspring. Resulting mutant cardiac phenotypes were confirmed through necropsy, or histopathologic analysis. Further analysis was completed examining the craniofacial and gross brain abnormalities discovered within the screen. Lines in which a mutant was recovered in the original screen were analyzed for both craniofacial and gross brain abnormality. Consistent phenotype in at least three mice was required for a line to be considered abnormal with a craniofacial or brain abnormality. An additional mouse model which recapitulates the phenotype of Joubert syndrome was analyzed prior to any analysis on the main *Ohia* line to validate method and findings. The mouse model was previously validated by Damerla et al.^73^ This Joubert syndrome mouse mode is colloquially called Heart Under Glass (*Hug*) and contains a S235P missense mutation in Jbts17. The *Hug* mice recapitulate the brain phenotype of Joubert syndrome presenting with decreased number of cerebellar fissures.

### Episcopic Confocal Microscopy (ECM)

Head samples were removed following necropsy and fixed in 4% PFA solution for a minimum of 72 hours. After fixation, samples were prepared for ECM via three graded ethanol baths of 10%, 20%, and 40% overnight at room temperature. Samples were then transferred to a Sakura Tissue Tek VIP 5 Tissue Processor where they were dehydrated using increasing percentages of warmed ethanol baths and eventually perfused with paraffin wax.

Samples were embedded in paraffin blocks to be sectioned coronally. 3D reconstructed images stacks were used for brain scoring and volumetric analysis. A diagram for the ECM workflow can be found in Supplemental Figure 4.

### Mouse Brain Dysplasia Scoring

A group of three reviewers reviewed 84 cases consisting of 69 Ohia, five CRISPR/CAS9, and 10 wild type samples. Reviewers came to a consensus for each structure scored. In total seven structures were included the mouse brain scoring including hippocampus, cerebellum, cerebrum, left and right olfactory bulbs, brain stem, and midbrain. Hippocampus, cerebellum, and both olfactory bulbs were scored for aplasia, hypoplasia, and dysplasia. Cerebrum, brain stem and midbrain were scored for hypoplasia and dysplasia only. Each instance of abnormality received a binary score of zero or one. Brain Dysplasia Score (BDS) was a sum of each area of the brain. Two other BDS metrics were calculated, one being a binary scoring of any cerebellar or hippocampal abnormalities and the another being a binary score if any abnormality was present in the sample at all. Cerebellar folds were counted using a sagittal view in the middle slice of the brain.

### Mouse Brain Segmentation

Tiff images were converted to NIFTI images using ITK-Snap and FSL. Images were down sampled 4x using FMIRB’s Linear Registration Tool (FLIRT). Resulting images were manually segmented into 16 discreet structures using ITK-Snap. Structures segmented included right hippocampus, right olfactory bulb, right subcortical area, right cortex, intracranial space, midbrain, left hippocampus, left olfactory bulb, left subcortical area, left cortex, extra-axial space, cerebellum, pons, medulla, hypothalamus, and choroid plexus. Due to the required processing methods for ECM, CSF is removed from the brain and replaced with paraffin wax. The space voided of CSF has been labeled as intracranial space and extra-axial space. Structures above the tentorium were combined to form a supratentorial volume and an infratentorial volume was also calculated. Infratentorial volumes consisted of the pons, medulla, and cerebellum, while the supratentorial volumes were composed of the hippocampus, hypothalamus, olfactory bulbs, subcortical areas, cortex, and midbrain.

### Genotype Analysis

We explored the relationship of Sap130/ Pcdha9 genotype and brain morphology in the mouse modeling. When comparing the risk stemming from the BDS for animals from the Ohia line, different genotypes were grouped into 6 distinct groups. Group A: *Sap130*(m/m) and *Pcdha9*(m/m), Group B: *Sap130*(m/m) and *Pcdha9*(m/+), Group C: *Sap130*(m/m) and *Pcdha9*(+/+), Group D: *Sap130*(m/+) and *Pcdha9*(m/m), Group E: *Sap130*(+/+) and *Pcdha9*(m/m), and lastly, Group F: *Sap130*(m/+) or (+/+) and *Pcdha9*(m/+) or (+/+).

### Mouse Genotype Groupings and Comparisons

For greater power in subsequent analysis, mice with similar genotypes were pooled into Groups. Group ABC consisted of mice which were homozygous mutant for *Sap130* (m/m), irrespective of their *Pcdha9* genotype. An additional Group, AB, contained mice which were homozygous mutant for *Sap130* (m/m), and either homozygous mutant or heterozygous for *Pcdha9* (m/*). To study the effect of homozygous mutations in the *Sap130* gene, Group ABC was compared against wildtype animals. Group ABC was then compared against Group F. Group F can only be heterozygous for *Sap130* and *Pcdha9*, so any effect of the homozygous mutant *Sap130* alleles would be removed in this group. This pairing should show effects of the homozygous *Sap130* mutation more concretely. To assess if any of the effect is coming from the *Pcdha9* genotype, Group AB was then compared to F. Group AB differs from Group ABC by removing samples that contained, wildtype *Pcdha9* genotyped animals. If *Pcdha9* is partially, or in large part, responsible for the observed phenotype, removing wildtype *Pcdha9* samples from the ABC Group should increase incidence and effects should become more significant. To further isolate the effect of *Sap130*, Group A was compared against Group C with the difference between groups being homozygous mutant *Pcdha9* (Group A) vs wildtype (Group C). Lastly, Group B was compared against Group D. Group B is homozygous *Sap130* and heterozygous *Pcdha9*, and Group D is the opposite, *Sap130* heterozygous, and *Pcdha9* homozygous mutant. If either homozygous gene is more significant than the other, the effect could be seen within this comparison.

### RNAseq Analysis

RNA was isolated from brain tissue samples of 4 *Ohia* mutant animals and 5 littermate controls at embryonic day E13.5-E14.5, libraries were constructed and sequenced on the Illumina HiSeq 2000 platform (BGI Americas), and differential gene expression analysis was conducted as previously described (Gabriel et al. Biorxiv paper). The online Database for Annotation, Visualization, and Integrated Discovery (DAVID) was used to perform gene ontology (GO) functional enrichment analysis.

### Statistical Analyses for Human Studies

#### Primary analysis

SAS statistical software was used to carry out human statistical analysis. Our primary analysis compared the derived BDS between neonates with CHD and controls. This analysis was performed using multivariate regression and three major co-variates were included in the model: gender, postconceptional age (PCA=gestational age plus postnatal age), time of MRI scan timing of scan relative to cardiac surgery (pre- vs post-operative). Multi-variate regression with false discovery rate (FDR) correction was used, to correct for multiple comparisons. The FDR is one way of conceptualizing the rate of type 1 errors in null hypothesis testing when conducting multiple comparisons. Whether BDS is association with sex difference, GA, or PCA was assessed in the entire study cohort.

#### Secondary analysis

Secondary analysis was repeated within the CHD only group, with birthweight and the presence of genetic abnormality included as additional comparisons with BDS. Furthermore, whether pre- or post-operative and pre-term or term status correlated with BDS was also analyzed within the CHD group. As an additional secondary analysis, within the CHD group, BDS was compared to the presence of various injuries and cortical maturation features. Additional secondary analyses including correlation of BDS with feeding data and neurodevelopmental outcome which consisted of linear and logistic regression with false discovery rate (FDR) correction for multiple comparisons; FDR-adjusted p-value <0.05 was considered significant.

### Statistical Analyses for Mouse Studies

Volumetric analysis of mouse individual structures was carried out using SAS statistical software. Both raw and normalized by total brain volume values were compared in any models generated. We completed an ad hoc analysis to delineate the relationship between BDS and regional brain volumes, clinical risk factor and heart lesion subtypes on our primary outcome measures. Clinical variables were compared using a three-way ANCOVA (analysis of covariance) between wild-type, *Ohia*, and *Hug* animals. We then compared each of the groups mentioned above to each other using pairwise T-tests.

## Results

### Comparison of Brain Injury/Cortical Maturation Score (TMS) between CHD and Control Cohorts

The preterm CHD and term CHD cohort demonstrated increased incidence of brain injury compared to their gestational-aged, matched controls. Specifically, the CHD preterm cohort demonstrated increased incidence of focal infarcts (p = 0.0295) and punctate white matter lesions (p = 0.0231) compared to the preterm control cohort. The term CHD cohort demonstrated increased rates of hemorrhage (p = 0.0284), focal infarct (p = 0.0035), punctate white matter lesions (p < 0.001), dichotomized injury composite score (p < 0.001), and injury composite score (p < 0.001) compared to the term control cohort (Supplemental Table 1).

When comparing the categories of the cortical maturation score (TMS), the term CHD cohort had widespread reduced cortical maturation (in all cortical lobes except occipital) compared to term control cohort, while the preterm CHD only differed from preterm controls in the myelination category (Supplemental Table 2).

### Derivation of Human BDS in Human Infant CHD

There was a high incidence of olfactory, hippocampal, and cerebellar abnormalities in the preterm and term CHD cohorts compared to both preterm and term control cohorts respectively (Table 1). Within the univariate analyses, there was no significant correlation between BDS and sex, GA, or PCA within the entire study cohort (Table 2A). Within the CHD cohort, there was no correlation between BDS and pre/postoperative status, preterm/term status, or birth weight. Higher BDS in the CHD cohort was correlated with known genetic abnormalities (Table 2B), which remained a significant association in the multivariable analysis (Table 2C). BDS was highly correlated with cortical immaturity by cortical maturation scores including frontal cortex (p < 0.0001), insular cortex (p < 0.0001), and cortical folding (p = 0.0016), even after controlling for PCA at the scan (Table 3). There was no significant correlation between BDS and any categories of brain injury (Table 3). Within a subset of cases the inter-rater reliability was calculated and resulted in reviewers one and two receiving high scores for most metrics included in the BDS calculation. Notable exceptions were for cortical folding (kappa = 0.18, 0.00), myelination (kappa = 0.35, 0.02), supratentorial extra-axial fluid (kappa = 0.35, 0.05), and germinal matrix (kappa = 0.48, 0.00). Other values for Cohen’s kappa measure for the BDS can be found in Supplemental Table 3.

**Table 1:** Incidence of Subcortical Brain Dysmaturation in Preterm/Term CHD And Controls.

| BDS Abnormalities (Dichotomous) | Preterm CHD<br>(n = 69) | Preterm Non-CHD (n = 51) | Comparison | Term CHD<br>(n = 263) | Term Control<br>(n = 93) | Comparison |
| --- | --- | --- | --- | --- | --- | --- |
|  | Number of Subjects with<br>Abnormality (%) |  | p-value | Number of Subjects with<br>Abnormality (%) |  | p-value |
| Bilateral Cerebellar Hemispheric Hypoplasia | 13 (18.84%) | 1 (1.96%) | **0.0049** | 30 (11.41%) | 0 (0.00%) | **0.0006** |
| Bilateral Cerebellar Hemispheric Dysplasia | 7 (10.14%) | 1 (1.96%) | 0.0607 | 15 (5.70%) | 0 (0.00%) | **0.0133** |
| Cerebellar Vermis Hypoplasia | 18 (26.09%) | 3 (5.88%) | **0.0040** | 58 (22.05%) | 0 (0.00%) | **<0.0001** |
| Cerebellar Vermis Dysplasia | 9 (13.04%) | 0 (0.00%) | **0.0049** | 23 (8.75%) | 0 (0.00%) | **0.0019** |
| *Dichotomized Cerebellar Composite* | 26 (37.68%) | 3 (5.88%) | **<0.0001** | 75 (28.52%) | 0 (0.00%) | **<0.0001** |
| Right Olfactory Bulb | 24 (34.78%) | 4 (7.84%) | **<0.0001** | 127 (48.29%) | 8 (8.60%) | **<0.0001** |
| Left Olfactory Bulb | 25 (36.23%) | 4 (7.84%) | **<0.0001** | 126 (47.91%) | 9 (9.68%) | **<0.0001** |
| Right Olfactory Sulcus | 23 (33.33%) | 1 (1.96%) | **<0.0001** | 109 (41.44%) | 3 (3.23%) | **<0.0001** |
| Left Olfactory Sulcus | 23 (33.33%) | 1 (1.96%) | **<0.0001** | 109 (41.44%) | 2 (2.15%) | **<0.0001** |
| *Dichotomized Olfactory Composite* | 28 (40.58%) | 4 (7.84%) | **<0.0001** | 127 (48.29%) | 9 (9.68%) | **<0.0001** |
| Hippocampus | 34 (49.28%) | 11 (21.57%) | **0.0019** | 117 (44.49%) | 3 (3.23%) | **<0.0001** |
| Corpus Callosum | 10 (14.49%) | 0 (0.00%) | **0.0045** | 26 (9.89%) | 0 (0.00%) | **0.0016** |
| Choroid Plexus | 30 (43.48%) | 3 (5.88%) | **<0.0001** | 108 (41.06%) | 8 (8.60%) | **<0.0001** |
| Brainstem | 10 (14.49%) | 0 (0.00%) | **0.0045** | 25 (9.51%) | 0 (0.00%) | **0.0020** |
| BDS Abnormalities (Categorical) | **Mean (Standard Error)** |  | **p-value** | **Mean (Standard Error)** |  | **p-value** |
| Supratentorial Extra-Axial Fluid* | 0.9839 (0.0989) | 0.6000 (0.1069) | **0.0075** | 0.8071<br>(0.0495) | 0.3516 (0.0527) | **<0.0001** |
| *Cerebellar Composite* | 0.6812 (0.1121) | 0.0980 (0.0578) | **<0.0001** | 0.4791<br>(0.0536) | 0.0000 (0.0000) | **<0.0001** |
| *Olfactory Composite* | 1.6912 (0.2693) | 0.1961 (0.1011) | **<0.0001** | 2.0229<br>(0.1452) | 0.2418 (0.0817) | **<0.0001** |
| *Subcortical BDS Composite* | 4.3188 (0.4043) | 1.1569 (0.2046) | **<0.0001** | 4.1141<br>(0.2210) | 0.6989 (0.1268) | **<0.0001** |

**Table 2A:** Correlation of Brain Dysplasia Score (BDS) with Demographic Factors (Entire cohort:Univariate Analysis)

| Dependent | Independent | N-used | R-Square | p-value | estimate |
| --- | --- | --- | --- | --- | --- |
| **BDS** | Sex | 490 | 0.000026 | 0.9096 | -0.0359 |
| **BDS** | GA | 487 | 0.003619 | 0.1851 | 0.058461 |
| **BDS** | PCA | 489 | 0.001664 | 0.368 | 0.011933 |

**Table 2B:** Correlation of Brain Dysplasia Score (BDS) with Demographic Factors (CHD cohort, Univariate Analysis)

| Dependent | Independent | N-used | R-Square | p-value | estimate |
| --- | --- | --- | --- | --- | --- |
| **BDS** | Sex | 344 | 0.008447 | 0.0887 | 0.680037 |
| **BDS** | Genetic Abnormality | 252 | 0.045926 | **0.0006** | -1.66277 |
| **BDS** | GA | 341 | 0.00002 | 0.9352 | -0.00562 |
| **BDS** | PCA | 343 | 0.002649 | 0.3419 | 0.017322 |
| **BDS** | Birth Weight | 248 | 0.000337 | 0.7737 | -9.1E-05 |
| **BDS** | Pre/Post operative status | 343 | 0.000662 | 0.6349 | -0.18132 |
| **BDS** | Term/Preterm | 343 | 0.000651 | 0.6378 | -0.23214 |

**Table 2C:** Correlation of Brain Dysplasia Score (BDS) with Demographic Factors in (CHD cohort, Multivariate Analysis)

|  |  |  | Independent |  | Term/Preterm |  | Pre/Post |  |
| --- | --- | --- | --- | --- | --- | --- | --- | --- |
| Dependent | N-used | R-Square | p-value | estimate | p-value | estimate | p-value | estimate |
| **Sex** | 343 | 0.009957 | 0.0854 | 0.6942 | 0.7409 | -0.1633 | 0.5872 | -0.2074 |
| **Genetic Abnormality** | 252 | 0.048769 | **0.0006** | -1.6806 | 0.3971 | -0.4741 | 0.8911 | -0.0617 |
| **GA** | 341 | 0.002196 | 0.6323 | 0.059 | 0.5401 | -0.5386 | 0.5487 | -0.2303 |
| **PCA** | 343 | 0.003471 | 0.3867 | 0.0168 | 0.6152 | -0.2489 | 0.8939 | -0.0543 |
| **Birth Weight** | 248 | 0.027317 | 0.4891 | 0.0002 | 0.0368 | -1.4958 | 0.0878 | -0.7169 |

**Table 3:**
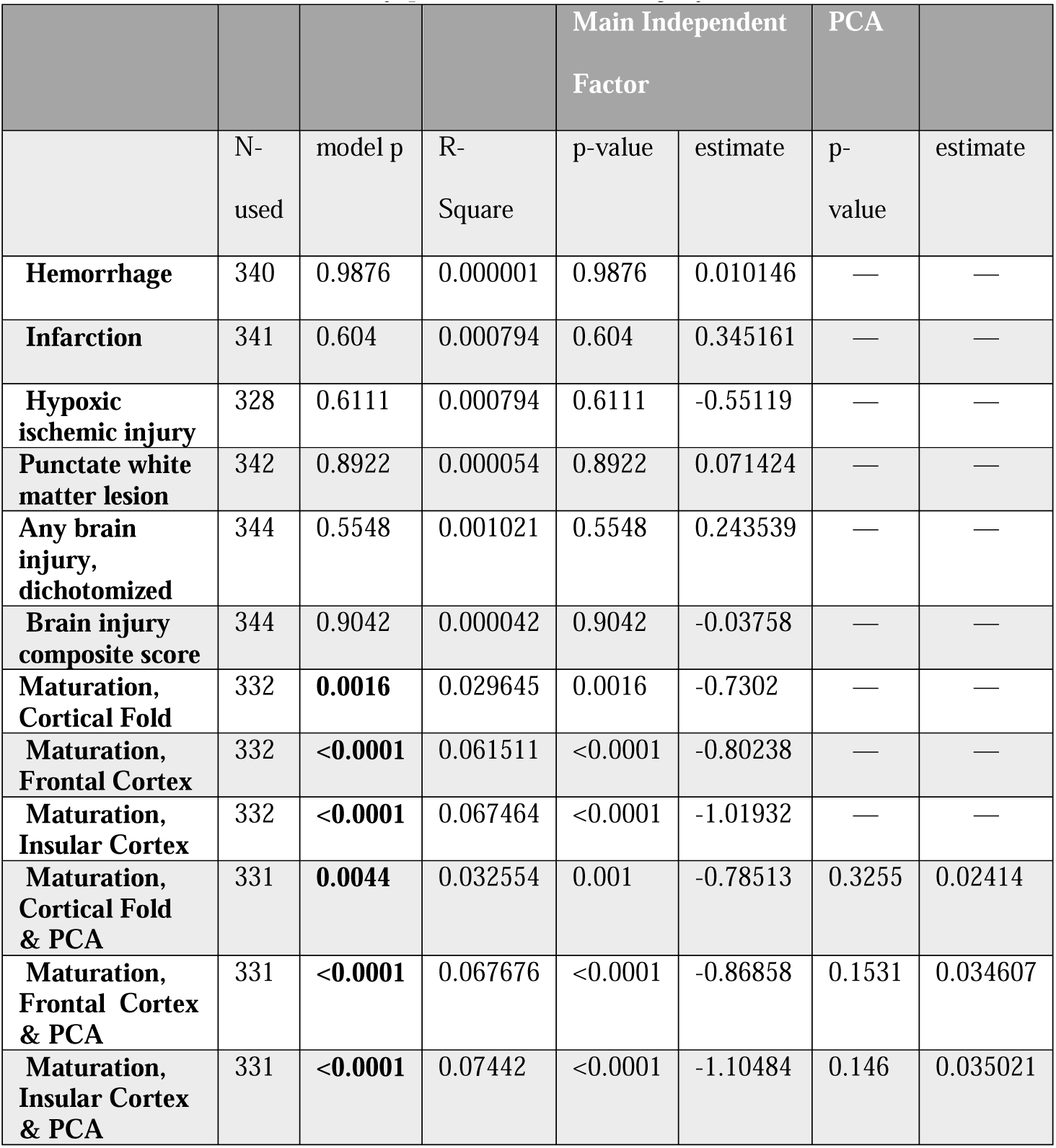
Correlation of Brain Dysplasia Score, Brain Injury and Cortical Maturation Score (TMS)

### Human Infant BDS and Regional Cerebral Volumes

Higher BDS correlated with smaller left and total cerebellar volume, smaller deep grey matter and brain stem volumes, and increased infra-ventricular and supra-tentorial CSF volumes in the entire sample (term CHD and term control cohorts combined) (Table 4). Increased BDS was more strongly correlated with reduced cerebellar volume, reduced cortical volumes, and increased CSF volume in the term CHD cohort compared to the term control cohort (Table 4).

**Table 4:** Association Between Brain Dysplasia Score and Term Infant Regional Brain Volumes by CHD Status.

| Normalized Volumes (*n* = 105) | Correlation with<br>BDS (term CHD<br>and term control<br>cohort combined) | Comparison between<br>term CHD and term<br>control cohorts |
| --- | --- | --- |
|  | **p-value**<br>**(*correlation*)** | **p-value (*direction*)** |
| Right Hippocampus | 0.0749 (*N.S.*) | 0.1476 (*N.S.*) |
| Left Hippocampus | 0.1329 (*N.S.*) | 0.3784 (*N.S.*) |
| Right Cerebellum | 0.0535 (*N.S.*) | 0.1476 (*N.S.*) |
| Left Cerebellum | **0.0433 (-)** | 0.1887 (*N.S.*) |
| Cerebellum | **0.0299 (-)** | **0.0087 (*CHD* > *CON*)** |
| Cerebrospinal Fluid (CSF) | 0.0517 (*N.S.*) | **0.0026 (*CHD* > *CON*)** |
| Infratentorial CSF | 0.2259 (*N.S.*) | 0.9300 (*N.S.*) |
| Intraventricular CSF | **<0.0001 (+)** | 0.7398 (*N.S.*) |
| Supratentorial CSF | **0.0008 (+)** | 0.3535 (*N.S.*) |
| Deep Grey Matter | **0.0138 (-)** | 0.4877 (*N.S.*) |
| White Matter | 0.4550 (*N.S.*) | 0.6066 (*N.S.*) |
| Cerebral Cortex | 0.1687 (*N.S.*) | **0.0016 (*CHD* > *CON*)** |
| Brainstem | **0.0391 (-)** | 0.1311 (*N.S.*) |

**Table 5:** Correlation between Brain Dysplasia Score and Clinical Feeding Outcomes.

| **BDS Abnormalities**<br>**(Clinical Outcome Correlates)** | **Lack of PO Feeds**<br>**Prior to**<br>**Discharge p-value** | **Length of**<br>**Hospitalization**<br>**p-value** |
| --- | --- | --- |
| **Hippocampus** | **0.0025** | **0.0090** |
| **Right / Left Olfactory Bulbs** | **0.0090 / 0.0050** | **0.0400 / 0.0300** |
| **Right / Left Olfactory Sulci** | **0.0070 / 0.0070** | **0.0700 / 0.0700** |
| **Extra-Axial Fluid** | **0.0030** | **<0.0001** |
| **Bilateral Cerebellar Hemispheres** | **0.0037** | **<0.0001** |
| **Cerebellar Vermis** | **0.0001** | **0.0002** |
| **Brainstem** | **<0.0001** | **N.S.** |
| **Corpus Callosum** | **0.0010** | **0.0100** |
| **Choroid Plexus** | **0.0100** | **0.0020** |
| ***Total Composite*** | **0.0001** | **0.0020** |

**Table 6:**
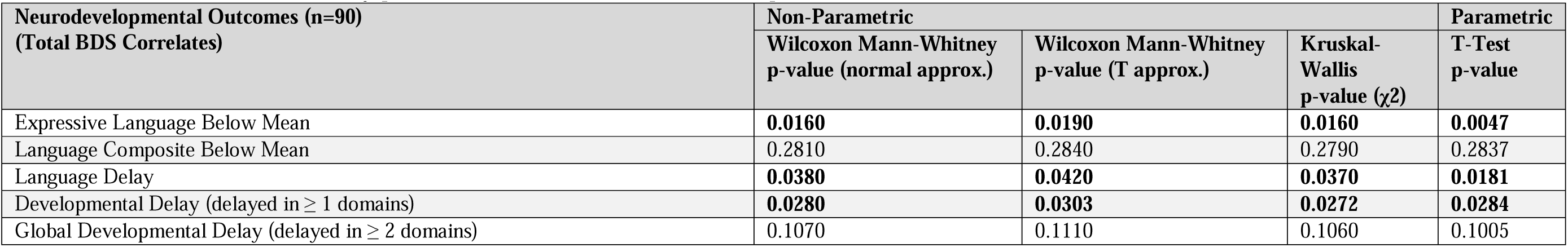
Correlation between Brain Dysplasia Score and 15-18 Month Neurodevelopmental Outcomes.

| Neurodevelopmental Outcomes (n=90)<br>(Total BDS Correlates) | Non-Parametric |  |  | Parametric |
| --- | --- | --- | --- | --- |
| | Wilcoxon Mann-Whitney<br>p-value (normal approx.) | Wilcoxon Mann-Whitney<br>p-value (T approx.) | Kruskal-Wallis<br>p-value ($\chi^2$) | T-Test<br>p-value |
| Expressive Language Below Mean | **0.0160** | **0.0190** | **0.0160** | **0.0047** |
| Language Composite Below Mean | 0.2810 | 0.2840 | 0.2790 | 0.2837 |
| Language Delay | **0.0380** | **0.0420** | **0.0370** | **0.0181** |
| Developmental Delay (delayed in $\geq 1$ domains) | **0.0280** | **0.0303** | **0.0272** | **0.0284** |
| Global Developmental Delay (delayed in $\geq 2$ domains) | 0.1070 | 0.1110 | 0.1060 | 0.1005 |

### Human Infant BDS and Clinical Risk Factors

When assessed against clinical risk factors in term CHD infants, the BDS did not correlate with birth factors, anthropomorphic data, cardiac lesion type, or intraoperative factors. (Supplemental Table 4A-D). The BDS did correlate with age of under 7 days at surgery (p<0.048, Supplemental Table 4B), increased length of hospitalization (p<0.0447, Supplemental Table 4D), and need for G-tube placement during the neonatal hospital stay (p<0.01, Supplemental Table 4D). Of note, we found no correlation between BDS and MRI field strength/scanner type (Supplemental Table 5).

### Human Infant BDS and Feeding Outcomes

Lack of oral feeding before neonatal hospital discharge was associated with multiple morphological abnormalities of the components of the BDS including cerebellar hemispheres and vermis, hippocampus, bilateral olfactory bulbs and olfactory sulci, corpus callosum, increased supratentorial extra-axial fluid, as well as total increased BDS. Hospital length of stay was associated with dysplasia of the cerebellar hemispheres and vermis, hippocampus, choroid plexus, brainstem, increased supratentorial axial fluid, and BDS. A diagnosis of dysphasia was associated with dysplasia of the brain stem (p= 0.001). Gastrointestinal dysmotility, aspiration, gastroesophageal reflux, malrotation, and vocal cord paralysis demonstrated no significant associations with brain dysplasia. Figure 2 panels A-D show some of the variations in hippocampal anatomy seen in the human population compared against that seen in the mouse in panels E and F. Figure 3 demonstrates olfactory bulb abnormalities in human infant CHD vs control (A/C) and the mouse model (B/D) . Figure 4 demonstrates cerebellar abnormalities in the human infant CHD vs controls (A-top) and the mouse model (A-bottom).

**Figure 2.**
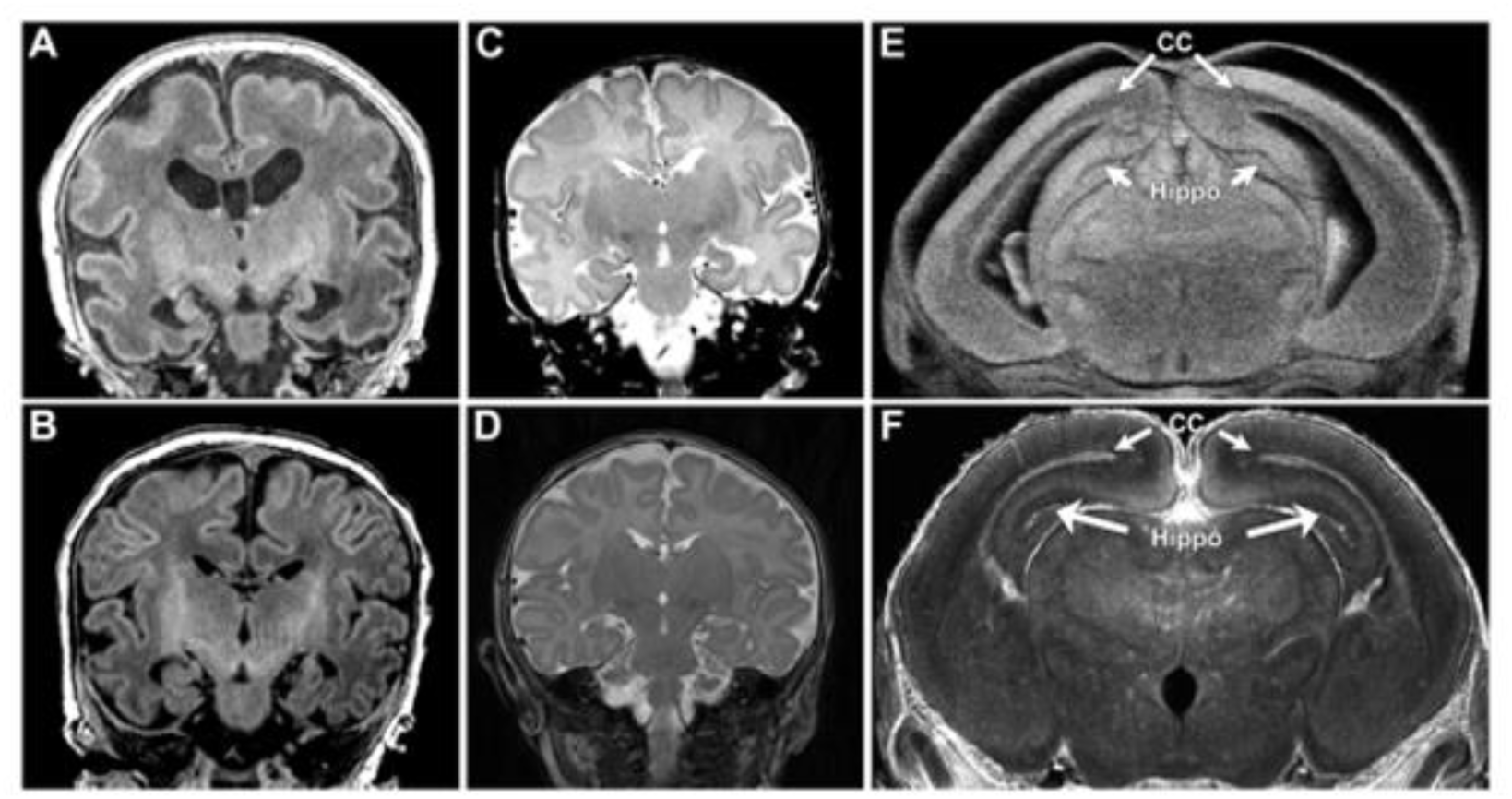
Hippocampal Abnormalities Were Similar Among Human and Mouse. Panel (a)(b)(c)and (d) demonstrate abnormal hypoplastic and/or malrotated hippocampi in infants with CHD which comparable abnormalities noted in the mouse mutant (e). A mouse control is shown for comparison in (f) depicts an abnormal. Panel (b)

**Figure 3.**
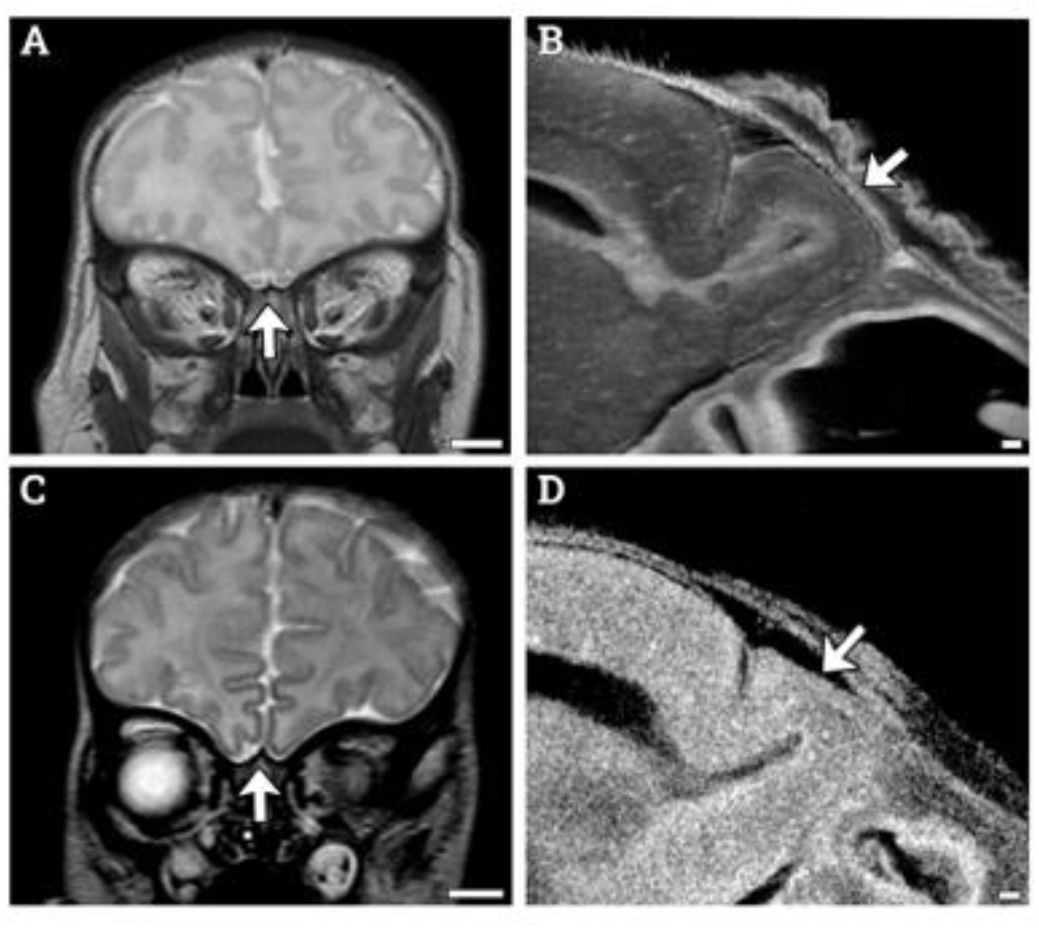
Consistent Olfactory Bulb Phenotype is Visible Among Human and Mouse. (a) Human control subject exhibits normal olfactory bulb size and structure. (b) Mouse control showing olfactory bulb with normal shape and size. (c) HLHS human subject with aplastic olfactory bulbs. (d) Mouse with Congenital heart disease with hypoplastic olfactory bulb.

**Figure 4.**
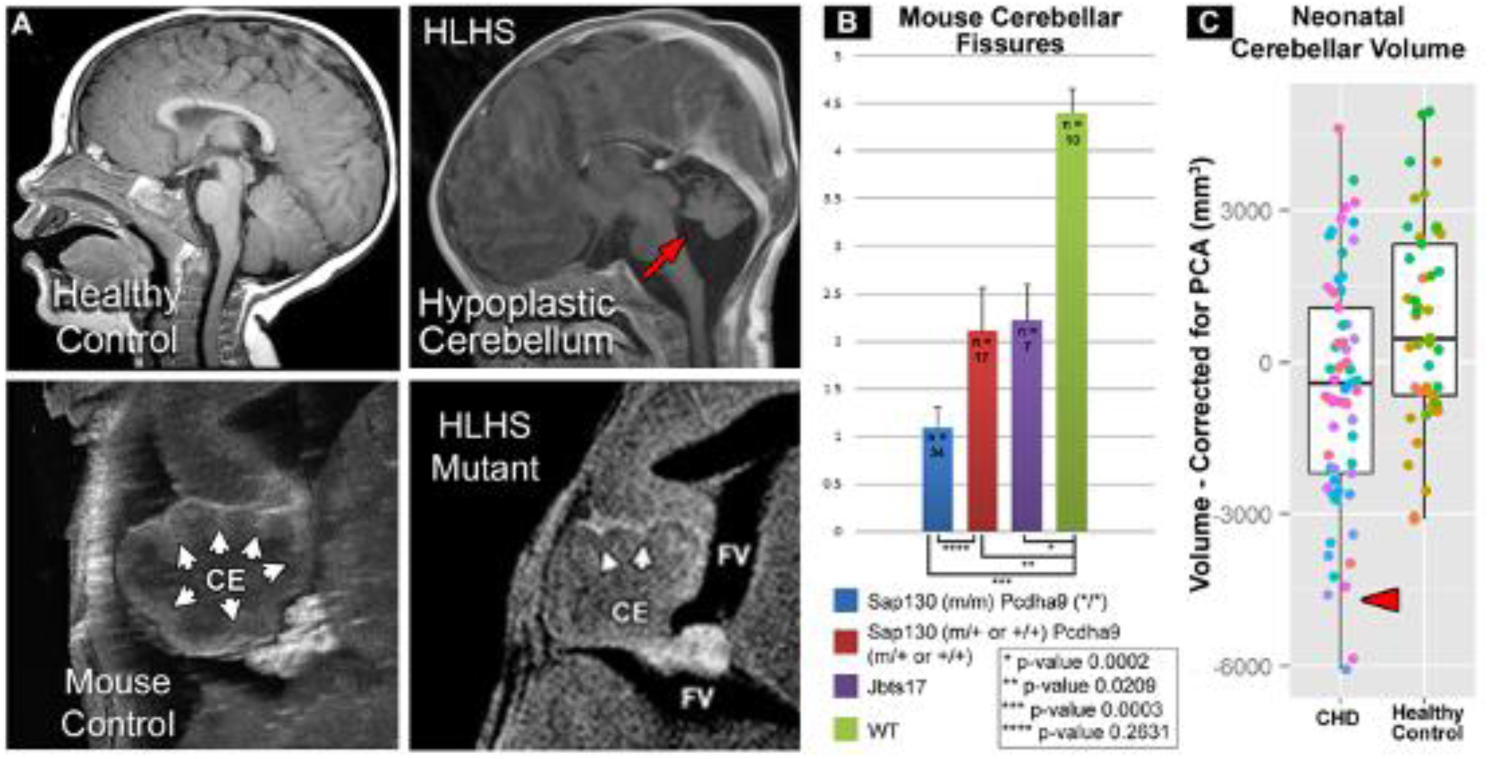
Cerebellar Abnormalities Seen in Both Human and Mouse. Panels (a) and (b) depict a human subject with HLHS compared to a healthy control. The cerebellum of the HLHS subject is hypoplastic shows dysmaturation. Panels (c) and (d) show a mouse with HLHS and a normal control. The cerebellum of the HLHS mutant displayed hypoplasia and a lesser number of cerebellar fissures. (e) Average number of cerebellar fissures among mouse groupings. (f) Human neonatal cerebellar residual volumes with gestational age regressed out.

### Human Infant BDS and Neurodevelopmental Outcomes

Increased BDS correlated with developmental delay in at least one domain including motor, cognitive, language/communication, or social/emotional (p<0.02), and specifically with poor expressive language development (p < 0.016) in a subset of the original sample tested at 15-18 months of age (n=90), given in Table 6.

### Mouse CHD Screen Results

From the original mouse screen for CHD, 3208 mouse pedigrees were screened, and 390 (12.16%) mutant lines were discovered. Of the 390 mutant lines, 214/390 (54.87%) had some form of craniofacial anomaly and 69/390 (17.69%) lines had a severe brain anomaly. Within those categories, craniofacial and brain anomalies, 174/214 (81.31%) and 62/69 (89.86%) had co-occurrence of CHD, respectively. Supplemental Figure 5 demonstrates some of the craniofacial abnormalities seen during the screening. 68 lines contained features of both types of anomalies, and of those 68 lines, 61 also exhibited CHD. A summary of the results can be seen in Figure 5

**Figure 5.**
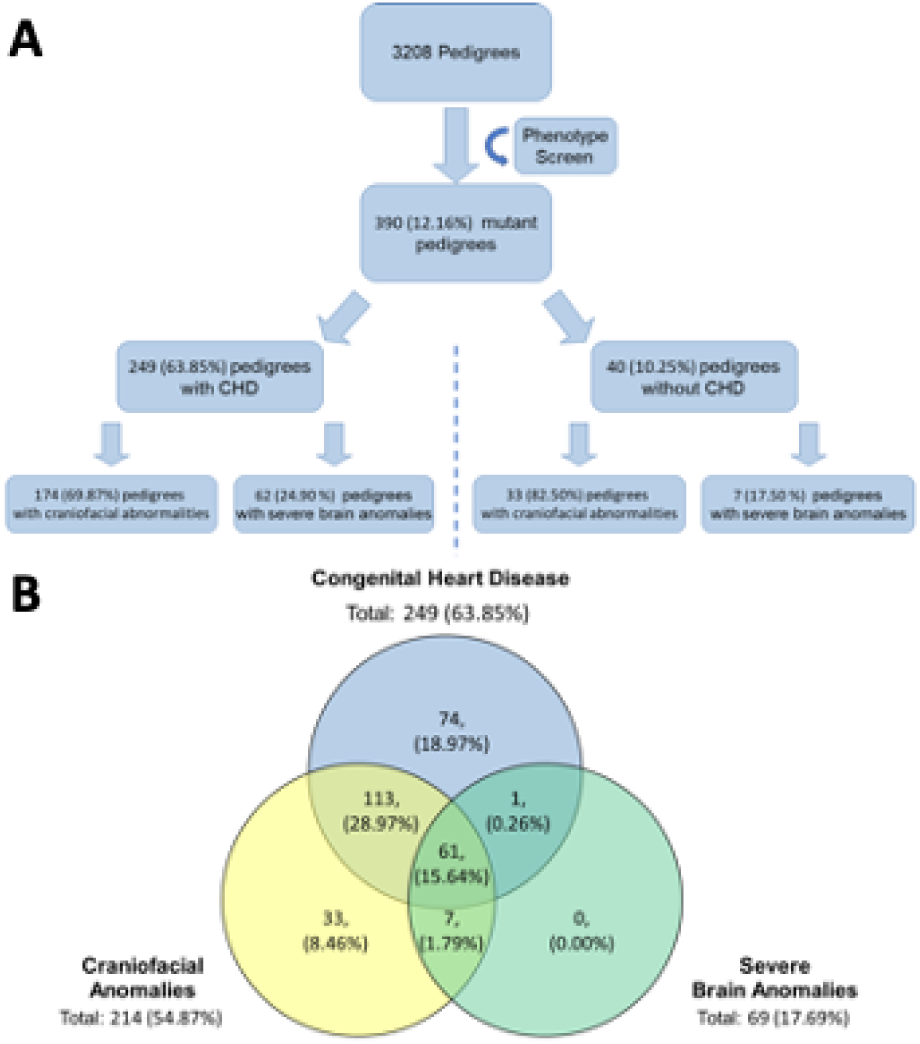
Phenotype Mouse Screen Recovered Mutant Mice with Defects in Cardiac, Brain and Craniofacial. (a) From the original phenotype screen 390 mutant lines were identified. Of those 390 lines 249 had CHD and of those, 174 and 62 had craniofacial and severe brain anomalies, respectively. Of the 40 lines that didn’t have CHD, 33 had craniofacial and 7 had severe brain anomalies. (b) Three major groupings, congenital heart disease, craniofacial anomalies, and server brain anomalies, identified from the original phenotype screen and their relationship to one another. The highest overlap between two conditions is congenital heart disease and craniofacial anomalies at approximately 29% of the original 390 mutants identified.

### Hug Model Mouse ECM Validation

To validate the ECM method, a mouse model, colloquially called Heart Under Glass or Hug, for Joubert syndrome with a S235P missense mutation in Jbts17 was analyzed prior to analysis of Ohia mice. Joubert syndrome is a ciliopathy associated with cerebellar abnormalities and other birth defects. In addition, Hug mice also had higher incidences of BDS (p =0.0161). and specifically, in the cerebellum-hippocampus dichotomized BDS (p =0.0154). Compared to wildtype mice, Hug mice showed an overall increase in cerebellar abnormalities (p < 0.001) and a decrease in the number of cerebellar fissures (p= 0.0023).

### Ohia Mouse Model Brain Dysplasia Scoring

ECM analysis was completed on 69 Ohia mutant neonatal mice. There was an increase in the BDS in the Ohia group compared to the wildtype group (p < 0.001) (Table 7). With regards to individual subcortical abnormalities (which make up the composition of the BDS), the cerebellum showed the highest incidence of abnormality with dysplasia being present in 49/67 (73.13%) of animals scored and was highly significant compared to control animals (p < 0.001). Ohia mutants also showed a decrease in the number of cerebellar fissures (p < 0.001) compared to the wild type group. The Ohia mutants also showed an increase in incidence of hippocampal dysplasia (p = 0.002), cerebral dysplasia (p = 0.003), left olfactory bulb aplasia (p = 0.024), left olfactory bulb any abnormality (p = 0.006), right olfactory bulb aplasia (p = 0.02), and right olfactory bulb any abnormality (p = 0.007) (Table 7). Supplemental Figure 6 demonstrates some of the common phenotypes seen in Ohia mice. Findings of aplasia, hypoplasia or dysplasia were common in the olfactory bulb, cerebellum, and/or hippocampus. Additionally holoprosencephaly was a coincidental finding in Ohia mice with 42/69 (60.9%) showing some signs of holoprosencephaly. Supplemental Figure 7 shows Ohia animals and two examples of the holoprosencephaly observed in this population.

**Table 7:** Incidence of Brain Dysplasia Score and Subcomponents between Ohia and Wild type Mouse Mutants.

| Structure | Ohia Average | Wild Type Average | P value † |
| --- | --- | --- | --- |
| **BDS Total** | **4.42** | 0 | **<0.001** |
| **Cerebellar Fissures** | **1.5** | 4.4 | **<0.001** |
|  | Ohia Incidence | Wild Type Incidence | P value * |
| **BDS Dichotomized** | **55/69, (79.71%)** | 0/10, (0.00%) | **<0.001** |
| **BDS Hippocampus or Cerebellum** | **58/69, (84.06%)** | 0/10, (0.00%) | **<0.001** |
| **Aplastic Hippocampus** | 8/69, (11.59%) | 0/10, (0.00%) | 0.384 |
| **Hypoplastic Hippocampus** | 14/68, (20.59%) | 0/10, (0.00%) | 0.223 |
| **Dysplastic Hippocampus** | **44/68, (64.71%)** | 0/10, (0.00%) | **0.002** |
| **Combination Hippocampus** | 22/69, (31.88%) | 0/10, (0.00%) | 0.133 |
| **Hypoplastic Cerebrum** | 13/68, (19.12%) | 0/10, (0.00%) | 0.244 |
| **Dysplastic Cerebrum** | **43/68, (63.24%)** | 0/10, (0.00%) | **0.003** |
| **Aplastic Cerebellum** | 2/69, (02.90%) | 0/10, (0.00%) | 0.678 |
| **Hypoplastic Cerebellum** | 9/67, (13.43%) | 0/10, (0.00%) | 0.345 |
| **Dysplastic Cerebellum** | **49/67, (73.13%)** | 0/10, (0.00%) | **<0.001** |
| **Combination Cerebellum** | 11/69, (15.94%) | 0/10, (0.00%) | 0.325 |
| **Aplastic Left Olfactory Bulb** | **33/69, (47.83%)** | 0/10, (0.00%) | **0.024** |
| **Aplastic Right Olfactory Bulb** | **34/69, (49.28%)** | 0/10, (0.00%) | **0.02** |
| **Hypoplastic Left Olfactory Bulb** | 9/67, (13.43%) | 0/10, (0.00%) | 0.345 |
| **Hypoplastic Right Olfactory Bulb** | 7/65, (10.77%) | 0/10, (0.00%) | 0.404 |
| **Dysplastic Left Olfactory Bulb** | 2/67, (2.99%) | 0/10, (0.00%) | 0.673 |
| **Dysplastic Right Olfactory Bulb** | 0/65, (00.00%) | 0/10, (0.00%) | — |
| **Combination Left Olfactory Bulb** | **42/69, (60.87%)** | 0/10, (0.00%) | **0.006** |
| **Combination Right Olfactory Bulb** | **41/69, (59.42%)** | 0/10, (0.00%) | **0.007** |
| **Hypoplastic Brainstem** | 0/66, (00.00%) | 0/10, (0.00%) | — |
| **Dysplastic Brainstem** | 20/67, (29.85%) | 0/10, (0.00%) | 0.12 |
| **Hypoplastic Midbrain** | 0/67, (00.00%) | 0/10, (0.00%) | — |
| **Dysplastic Midbrain** | 18/67, (26.87%) | 0/10, (0.00%) | 0.148 |

† Student's T-test was used to compare Cerebellar Fissure count and BDS Total Score.\* Chi-square analysis was used to compare individual structures as well as BDS Dichotomized and BDS Hippocampus or Cerebellum.

### Ohia Brain Dysplasia Scoring within CHD Subgroupings

To examine the relationship between CHD lesions and brain dysmaturation, the co-occurrence of different brain anomalies and any form of CHD was compared. We compared within the Ohia cohort looking at BDS and cerebellar fissures and found that the presence of CHD increased the BDS and reduced cerebellar fissures (p- < 0.001) (Table 8). Looking at other forms of cardiac sub-lesions, there was no significant differences between BDS and cerebellar folds (Table 8).

**Table 8:** Relationship between Brain Dysplasia Scores/Cerebellar Fissures and Cardiac Lesion Subtypes in Ohia CHD/HLHS Preclinical Groups.

|  | CHD vs. No CHD |  |  | Single vs. Biventricular Morphology |  |  | Conotruncal vs. Non Conotruncal |  |  | Cyanotic vs. Acyanotic |  |  | Arch Obstruction vs. No Obstruction |  |  |
| --- | --- | --- | --- | --- | --- | --- | --- | --- | --- | --- | --- | --- | --- | --- | --- |
|  | CHD | No CHD | P value * | Single | Double | P value * | Conotruncal | Non Conotruncal | P value * | Cyanotic | Acyanotic | P value * | Obstruction | No Obstruction | P value * |
| Cerebellar Fissures | 1.17 ± 1.55 | 3.14 ± 1.67 | **<0.001** | 1.50 ± 2.12 | 1.16 ± 1.55 | 0.86 | 1.13 ± 1.92 | 1.22 ± 1.36 | 0.88 | 1.16 ± 1.80 | 1.19 ± 1.39 | 0.96 | 0.81 ± 0.98 | 1.48 ± 1.87 | 0.13 |
| BDS Total | 5.43 ± 2.22 | 2.73 ± 2.46 | **<0.001** | 7.00 ± 2.00 | 5.32 ± 2.20 | 0.21 | 5.27 ± 2.12 | 5.33 ± 2.46 | 0.93 | 5.63 ± 2.34 | 5.29 ± 2.17 | 0.61 | 5.87 ± 1.69 | 5.00 ± 2.60 | 0.18 |

\* Student T-test used to calculate difference between population values within cerebellar fissures and Total Brain Dysmaturation Score (BDS).

In Ohia animals there were 10 brain structures which showed an increased incidence of abnormalities in animals with CHD. Those structures included hippocampal dysplasia (p = 0.043), cerebral hypoplasia (p = 0.008), cerebral dysplasia (p < 0.001), cerebellar dysplasia (p < 0.001), left olfactory bulb aplasia (p < 0.001), left olfactory bulb any abnormality (p < 0.001), right olfactory bulb aplasia (p < 0.001), right olfactory bulb any abnormality (p < 0.001), brainstem dysplasia (p = 0.016), and midbrain dysplasia (p = 0.0062) (Supplemental Table 6A). Comparing single versus biventricular morphology, defects were more common in the cerebellum in double ventricular mice (p < 0.001) (Supplemental Table 6A). When comparing conotruncal vs non-conotruncal defects, mice with conotruncal defects showed an increased in the occurrence of hypoplasia in the cerebrum (p = 0.037) (Supplemental Table 6B) Within cyanotic vs acyanotic cardiac defects, mice with acyanotic defects showed increases in the incidence of left olfactory bulb hypoplasia (p = 0.018) and any defect in the cerebellum (p = 0.037). Lastly, looking at mice with and without arch obstructions, mice with arch obstruction showed an increase in the occurrence of any defect in the cerebellum (p = 0.048) (Supplemental Table 6B).

### Ohia Mouse Model Regional Brain Volumes

Ohia mutants demonstrated a significant decrease in raw (non-normalized) volumes of the right subcortical (p = 0.044), left subcortical (p = 0.041), cerebellar (p = 0.005) and supratentorial volume (p = 0.037) volumes (non-normalized) compared to wild type control. Ohia mutants also demonstrated an increase in the intraventricular volume (p = 0.009) compared to wild type controls (Supplemental Table 7.

Normalizing volumes to total brain volume showed Ohia animals also exhibited a significant increase in medullary (p = 0.024) and intraventricular normalized volume (normalized to total brain volumes). (p = 0.015). Ohia animals also demonstrated a decrease in normalized supratentorial volumes (p = 0.01) compared to wild type controls (Supplemental Table 7).

### Ohia Mouse Model Regional Brain Volumes within CHD Subgroupings

There were no differences in raw (non-normalized) volumes between Ohia mice with and without CHD. Looking at single versus biventricular morphology, mice with biventricular heart defects showed increases in the volumes of the right subcortical areas (p = 0.006), left subcortical area (p = 0.036), and the pons (p = 0.024) (Supplemental Table 8a). Comparing conotruncal against non-conotruncal heart defects, mice with conotruncal defects showed an increased in the right subcortical area (p = 0.043) (Supplemental Table 8b). Mice with acyanotic defects demonstrated an increase in the volume of the left hippocampus (p = 0.044) (Supplemental Table 8b). There were no differences in volume noticed between mice with and without arch obstructions.

After normalizing raw volumes to total brain volume different regions showed significance between groups. Between CHD and non-CHD Ohia mice, Ohia mice with CHD showed increased volumes in the right cortex (p = 0.026), midbrain (p = 0.044) and cerebellum (p = 0.044). Comparing normalized volumes within single and biventricular groupings found that single ventricular morphology heart defect mice had increased choroid plexus volumes (p < 0.001). (Supplemental Table 9a). There were no significant findings looking between groups of conotruncal vs non-conotruncal, cyanotic vs acyanotic, or arch obstruction vs no arch obstruction (Supplemental Table 9b).

### Ohia Mouse Model Brain Dysplasia Scoring within Genotype Subgroupings

Comparing genotypes within the Ohia cohort, samples were first grouped based on genotypes. The two groups with the most samples were group A which consisted of Sap130 and Pcdha9 double mutants and group F consisted of animals which did not have a Sap130 or Pcdha9 homozygous mutant genotype (Table 9). Both groups showed high incidence rates of olfactory bulb, cerebellar, hippocampal, and brainstem abnormalities (Table 9). Using Group ABC mentioned in the Mouse Genotype Groupings section in the methods, values of hippocampus and cerebellar BDS (p < 0.001) and BDS total (all structures) were greater (p < 0.001) in Group ABC compared to wildtype (Table 10).

**Table 9:** Relationship of Genotype to Incidence of Subcortical Abnormalities used to Derive BDS score in CHD/HLHS Preclinical Groups.

| Group | A<sup>†</sup> | B<sup>‡</sup> | C<sup>§</sup> | D | E<sup>¶</sup> | F<sup>#</sup> |
| --- | --- | --- | --- | --- | --- | --- |
| **N Total** | 24 | 8 | 7 | 4 | 0 | 20 |
| **Olfactory bulb abnormality** | 18 (75%) | 6 (75%) | 6 (85.7%) | 4 (100%) | 0 | 9 (45%) |
| **Cerebellar abnormality** | 21 (87.5%) | 7 (87.5%) | 7 (100%) | 4 (100%) | 0 | 13 (65%) |
| **Hippocampus abnormality** | 21 (87.5%) | 7 (87.5%) | 6 (85.7%) | 4 (100%) | 0 | 13 (65%) |
| **Brainstem abnormality** | 10 (42%) | 2 (25%) | 0 (0%) | 2 (50%) | 0 | 6 (30%) |

†Group A: N 24, Genotype: Pcdha9(m/m) Sap130(m/m)
‡ Group B: N 8, Genotype: Pcdha9(m/+) Sap130(m/m)
§ Group C: N 7, Genotype: Pcdha9(+/+) Sap130(m/m)
|| Group D: N 4, Genotype: Pcdha9(m/m) Sap130(m/+)
¶ Group E: N 0, Genotype: Pcdha9(m/m) Sap130(+/+)

**Table 10:** Relationship of Genotype to BDS Score in CHD/HLHS Preclinical Groups.

|  | **ABC\*\* vs. WT<sup>‡‡</sup>** |  |  | **ABC\*\* vs. F<sup>#</sup>** |  |  | **AB<sup>††</sup> vs. F<sup>#</sup>** |  |  | **A<sup>†</sup> vs. C<sup>§</sup>** |  |  | **B<sup>‡</sup> vs. D** |  |  |
| --- | --- | --- | --- | --- | --- | --- | --- | --- | --- | --- | --- | --- | --- | --- | --- |
| **Structure** | ABC<br>Median | WT<br>Median | P value* | ABC<br>Median | F<br>Median | P<br>value * | AB<br>Median | F<br>Median | P<br>value * | A<br>Median | C<br>Median | P<br>value * | B<br>Median | D<br>Median | P<br>value * |
| **BDS**<br>**Hippocampus**<br>**or**<br>**Cerebellum** | 1 | 0 | **<0.001** | 1 | 1 | **0.008** | 1 | 1 | **0.021** | 1 | 1 | 0.583 | 1 | 1 | 0.46 |
| **BDS Total** | 6 | 0 | **<0.001** | 6 | 5.5 | 0.227 | 6 | 5.5 | 0.251 | 6 | 5 | 0.56 | 6 | 6.5 | 0.558 |

\* Non-parametric Mann–Whitney U test used to analyze differences in continuous Brain Dysmaturation Score.
†Group A: N 24, Genotype: Pcdha9(m/m) Sap130(m/m)
‡ Group B: N 8, Genotype: Pcdha9(m/+) Sap130(m/m)
§ Group C: N 7, Genotype: Pcdha9(+/+) Sap130(m/m)
|| Group D: N 4, Genotype: Pcdha9(m/m) Sap130(m/+)
¶ Group E: N 0, Genotype: Pcdha9(m/m) Sap130(+/+)

Within the different genotype groups there were various findings. Groups A and B had 19/24 (79.17%) and 6/8 (75%) of mice exhibiting some form of holoprosencephaly. Additionally, those groups had the high rates of CHD and the various subtypes of CHD (i.e., conotruncal, single ventricular morphology, cyanotic defects, and arch obstructions) (Supplemental Table 10.

When comparing ABC vs WT, to determine homozygous Sap130 mutation difference compared to controls, 10 structures studied showed greater incidence values in ABC compared to wildtype. Those structures included dysplastic hippocampus (p < 0.001), hypoplastic cerebrum (p = 0.40), dysplastic cerebrum (p < 0.001), dysplastic cerebellum (p < 0.001), aplastic left olfactory bulb (p = 0.001), combination of left olfactory bulb (p < 0.001), aplastic right olfactory bulb (p = 0.001), combination of right olfactory bulb (p = <0.001), dysplastic brainstem (p = 0.037), and the dichotomized BDS (p = <0.001) (Supplemental Table 7a).

Comparing groups ABC vs. F to determine homozygous Sap130 mutation difference compared to no homozygous mutation in either gene (at most hetero or WT for either mutation), hippocampal and cerebellar BDS was different between groups with ABC, a higher incidence (p = 0.008) (Table 10). Eight structures had significantly increased incidence in group ABC compared to F. Structures included hypoplastic cerebrum (p = 0.021), dysplastic cerebrum (p = 0.016), dysplastic cerebellum (p < 0.001), aplastic left olfactory bulb (p = 0.035), combination of left olfactory bulb (p = 0.010), aplastic right olfactory bulb (p = 0.022), combination of right olfactory bulb (p = 0.018), and dichotomized BDS (p = 0.021). Additionally, the ABC group had an increased value for the hippocampus and cerebellum BDS (p = 0.008) (Supplemental Table 7a).

In comparing groups AB vs. F, (homozygous Sap 130 mutation and homo/heteroPCDHA9 vs no hippocampal and cerebellar BDS was significantly different between groups (p 0.021) (Table 10). Seven structures showed significant differences in the rate of abnormalities; structures included hypoplastic cerebrum (p = 0.035), dysplastic cerebrum (p = 0.019), dysplastic cerebellum (p = 0.001), aplastic left olfactory bulb (p-value = 0.039), combination of left olfactory bulb (p = 0.023), aplastic right olfactory bulb (p = 0.023), and combination of right olfactory bulb (p = 0.023) (Supplemental Table 7b). Comparing groups A vs C, only 2 structures were found significant. Those structures were dysplastic brainstem (p = 0.028) and dysplastic midbrain. (p = 0.042) (Supplemental Table 7b).

Comparing groups B vs D, only aplastic hippocampus (p 0.029) and any defect in hippocampus (p = 0.014) were found to be significant (Supplemental Table 7c).

### Ohia Mouse Model Regional Brain Volumes within Genotype Subgroupings

Looking at volumes for brain regions between genotype groups, in group ABC vs wild type, six areas were found to be significantly different with mice in group ABC having reduced volumes including the right subcortical area (p = 0.006), right cortex (p = 0.047), left subcortical area (p = 0.007), cerebellum (p-value = 0.003), supratentorial volume (p =0.013), and total brain volume (p = 0.028) (Supplemental Table 11a).

In group ABC vs F, mice in group ABC showed significant volume reductions in eight structures. Structures included right hippocampus (p-value = 0.041), right subcortical areas (p = 0.050), right cortex (p = 0.016), left hippocampus (p-value = 0.031), left olfactory bulb (p = 0.048), left cortex (p = 0.022), supratentorial volume (p-value = 0.018), and total brain volume (p = 0.022) (Supplemental Table 11a). Similarly comparing groups AB vs. F, mice in group AB showed significant reductions in volumes in eight structures. Differences occurred between right hippocampus (= 0.021), right subcortical areas (p-value = 0.023), right cortex (p = 0.029), left hippocampus (p-value = 0.024), left subcortical areas (p =0.038), left cortex (p =0.029), supratentorial volume (p =0.014) and total brain volume (p = 0.014) (Supplemental Table 11b). Lastly, comparing group A vs C, animals in group A showed significant reductions in four areas including the right subcortical area (p =0.022), left subcortical area (p =0.034), infratentorial volume (p =0.047), total brain volume (p =0.049) (Supplemental Table 11b).

After normalizing raw volumes to total brain volume, multiple brain regions showed significant differences between groupings. The ABC group demonstrated increased volume of the midbrain (p = 0.027) and medulla (p = 0.005) and decreased volume of the right olfactory bulb (p = 0.037) and supratentorial region (p = 0.009) compared to the wild type groups (Supplemental Table 12a). The ABC group demonstrated significant increases in volume in three areas including the cerebellum (p = 0.010), medulla (p = 0.043), and choroid plexus (p = 0.012) compared to the F group. Additionally, the right cortex volume was reduced in animals in group ABC compared to F (p = 0.028) (Supplemental Table 12a).

Comparing groups AB vs F, mice in group AB demonstrated three structures with significant increases in volume including the cerebellum (p = 0.012), pons (p = 0.049), and choroid plexus (p = 0.029). There were no significant differences between groups A vs C and B vs D (Supplemental Table 12b).

### Mouse CHD/Ohia RNA-seq

RNA-seq transcriptome profiling in term Ohia brains showed changes in genes involved in learning and memory, feeding behaviors, neuronal differentiation, and synaptic transport (Figure 6) Sap130 chromatin immunoprecipitation sequencing (CHIP-seq) looking at targets associated with Sap130 in brain tissue revealed a total of 644 genes being associated with cardiac and brain associated genes. Of those 644 genes, 106 and 324 were cardiac and brain genes, respectively, with the remaining 214 genes being associated with both brain and cardiac. The top three phenotypes associated with the Sap130 CHIP-seq analysis were global brain delay, abnormal brain development, and abnormal nervous system development.

**Figure 6.**
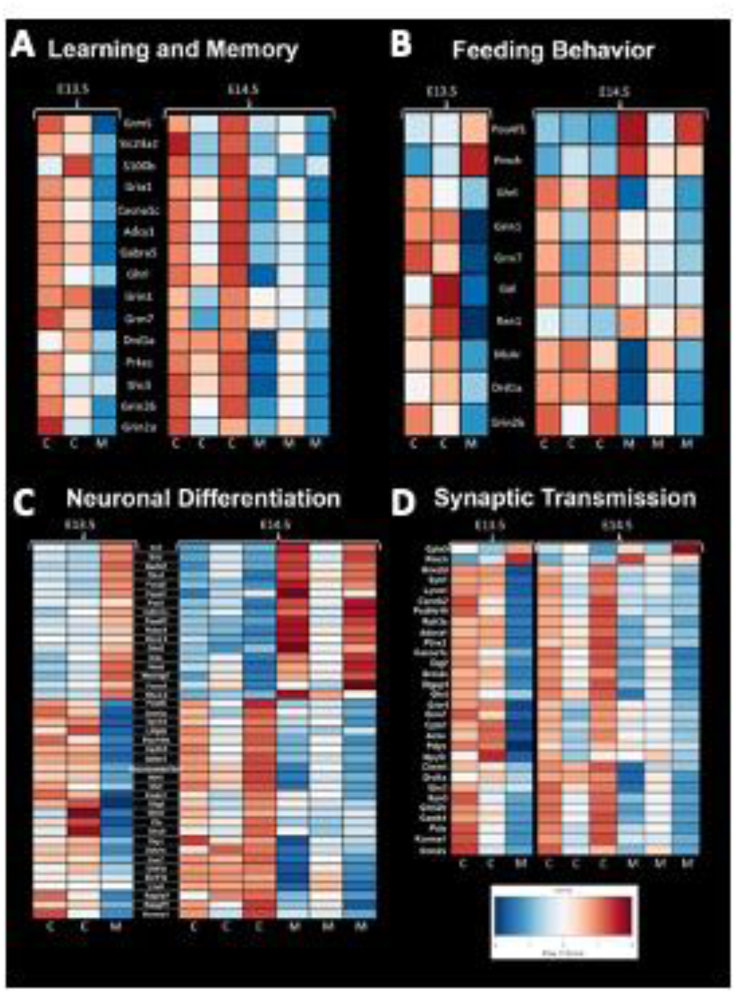
Mouse RNA Sequencing Analysis Shows Significant Difference in Gene Expression. (a) Genes associated with learning and memory showed lesser expression in control mice compared to Ohia mutant mice. (b) Genes associated with feeding behavior showed an abnormal pattern in Ohia mice. Ohia mice showed a pattern of expression that was opposite compared control mice at both time points. (c) Genes associated with neuronal differentiation showed a opposite pattern of expression in Ohia mutant mice compared to controls. (d) Genes associated with synaptic transmission had a different expression pattern in Ohia mutant mice compared to controls.

### Crisper/CAS Phenotype Validation

Crisper/CAS9 mice were also processed via ECM for validation of phenotype associated with Ohia mice. Three different crisper/CAS mice were developed to validate, a Sap130/Pcdha9 double mutant, Sap130 only and Pchda9 only. Brain scoring was performed like Ohia mice and findings support a similar phenotype. In the Sap130/Pcdha9 double mutant mice analyzed, they were found to exhibit a severe phenotype characterized by smoothed cerebellum, aplastic or highly dysplastic olfactory bulbs, and aplastic hippocampus. Other animals showed a pattern of holoprosencephaly. The Sap130 crisper animal showed a severe phenotype as well with smooth cerebellum, absent or extremely abnormal olfactory bulbs, and either holoprosencephaly or an abnormal hippocampus. In the Pcdha9 crisper animal, the phenotype was milder and presented with a cerebellum that while dysplastic did present with fissures unlike the double mutant or Sap130 crisper animal, a semi-normal appearing olfactory bulb and hippocampus. The phenotype was more severe in animals having mutations in the Sap130 gene. A visual representation of the defects found in Crisper/CAS9 mice can be found in Supplemental Figure 8. A similar pattern of severe olfactory bulb and hippocampal structural abnormalities was seen in our human population. Supplemental Figure 9 shows the Crisper/CAS animals compared to a human subject with severe brain dysmaturation.

## Discussion

Our study validates a paralimbic related subcortical based semi-quantitative BDS, which has proven to be sensitive to dysmaturational differences between CHD and control infants as well as sensitive to neurodevelopmental outcomes within CHD infants, including associations with early language, clinical and feeding outcomes. Most conventional semi-quantitative MRI scoring systems in CHD patients have exclusively focused on either acquired brain injury or cortical maturation assessment, particularly in the neonatal period.^26–33^ In this study, we demonstrated that not only is it possible to generate a paralimbic-related subcortically informed scoring system, but that such a scoring system holds important information regarding outcomes for CHD infants at risk for atypical neurodevelopment. The key subcortical components of our scoring system included subcortical structures that are known to be part of para-limbic neural networks including the olfactory bulb, the hippocampus, the cerebellum and the deep grey structures like the striatum. Furthermore, we showed that there is a distinct similarity between findings seen in CHD infants when compared to a preclinical mouse model of CHD suggesting similar mechanisms of brain dysmaturation.

In our study, we found a high incidence of paralimbic related subcortical abnormalities including olfactory, cerebellar, hippocampal, and brainstem abnormalities in infants with CHD compared to controls. We found similar olfactory bulb defects in the preclinical mouse model that we also see in our human CHD population. These abnormalities were used to derive a semi-quantitative subcortical abnormality or brain dysplasia score (BDS). We found that this BDS correlated with reduced cortical maturation (measured with the classic TMS), increased CSF volume and increased deep grey volume (striatum/thalamus). In contrast, the BDS did not correlate with GA, preterm status, cardiac lesion subtype, birth factors, intraoperative factors, or different vendor MRI/field strength. BDS did correlate with developmental delay and poor expressive language development. The BDS also correlates with age at surgery, increased length of hospitalization and need for G-tube placement. A more detailed evaluation of factors surrounding feeding and individual brain dysplasia parameters found correlations between lack oral feeding prior to neonatal hospital discharge and dysplasia of multiple brain areas as well as the BDS, and additionally with hospital length of stay which is likely the sequala of feeding dysfunction. Interestingly, swallowing dysfunction was associated with dysplasia of the brain stem.

An important component of our paralimbic-related subcortical BDS scoring system is the hippocampus. We found a consistent pattern of dysmaturation in the hippocampus, including severe forms of hippocampal aplasia or dysplasia in both mice and human subjects. Research has indicated that reductions in hippocampal volume and functional correlates in adolescents and adults resulted in an increased rate of neurodevelopmental impairment ^74,75^. Fontes et. al found that hippocampal shape and volumetric abnormalities found within CHD subjects can be predictive of impaired executive function. It is possible that the hippocampus is particularly vulnerable to early injury in CHD individuals as it is a brain region particularly susceptible to injury related to hypoxemia or hemodynamic instability. Latal et al found that reductions in hippocampal volume correlated with reductions in total IQ, working memory, and verbal comprehension. Lastly, one study looking at maternal stress found that reductions in hippocampal volumes are present in utero, consistent with previous volumetric findings of CHD^76^. Wu et. al concluded that universal screening looking at maternal stress is important, as early identification of hippocampal abnormalities is imperative.

Another important component of our paralimbic-related subcortical scoring system was the cerebellum. Cerebellar dysfunction has been linked with adverse neurodevelopmental outcomes^77–79^. Stoodley found that cerebellar damage or malformation present in early stages of development to be more detrimental than when obtained in adulthood, and theorized that injuries could affect cerebral-cerebellar circuits that are crucial to development and learning.^78^ Zwicker et. al found that preterm infants with exposure to morphine in the neonatal period had decreased cerebellar volumes and worse neurodevelopmental outcomes.^79^ Additionally, our group has previously shown that CHD children with reduced cerebellar volumes scored worse in tests for working memory, inhibitory control, and mental flexibility^80^. We also demonstrated that the superior surface of the cerebellum, primarily composed of the posterior lobe and the midline vermis, is an area particularly susceptible to alterations in morphology, indicating possible regions specifically affected by dysmaturation in CHD.^35^ Previous literature in combination with our BDS findings show the importance of the cerebellum toward neurodevelopment and suggest that insults or injury affecting proper cerebellar development may impact neurodevelopmental outcomes. Figure 4 demonstrates some of the commonalities seen between the mouse and human cerebellum; of note are the abnormal number cerebellar fissure and small size.

An important subcortical structure that was noted to be abnormal in the human BDS score volumetric correlation analysis and in the preclinical modeling was the striatum. In infancy, striatal connectivity is related to later motor and affective outcomes^81–83^, with previous research suggesting there are abnormalities in CHD^84^, potentially linking disrupted development with future behavior. The striatum forms a neural network with indirect and direct connectivity with other subcortical paralimbic components of the BDS score including the cerebellum, hippocampus and olfactory bulb. As such the developing striatum is part of a dense, interconnected network that supports motor function, reward processing, and cognition^85,86^, with circuitry specialized for affective based learning and olfactory reward. Supported by work in animal models and adults, the olfactory bulb directly projects to the olfactory tubercle^87^, a part of the ventral striatum, specialized for olfaction and containing significant dopamine neurons^88,89^. Together with innervations to the striatum from the hippocampus and amygdala^90–93^, the ventral striatum has been suggested to represent odor valence cues and instigate goal-directed behavior^87^, facilitated by additional input from piriform cortex and prefrontal cortex^94–96^. Output from the ventral striatum proceeds to the pallidum and then the thalamus, which then closes the feedback loop with prefrontal cortex^97,98^. An important structure in this loop is the orbitofrontal cortex, which plays an important role in maintaining changing odor representations but also more broadly in decision making and stimulus evaluation^99^. Integrated into these loops are other subcortical structures, including the cerebellum, which also has a significant role in movement and more recently emphasized role in cognition and language^100^. Through the thalamus and pons, the cerebellum indirectly projects to putamen, pallidum and the subthalamic nucleus^101,102^. These outputs may be topographically organized, converging on both the ventral striatum and frontal areas, with suggested involvement in appetitive reward^103^ and strategic motor learning^104^. Disruption of this interconnected network is likely captured by BDS scores, which are then sensitive to striatally relevant neurodevelopmental outcomes.

Within the *Ohia* mouse model the two main driver genes were *Sap130* and *Pcdha9*, and while both genes were necessarily to have cardiac phenotype, both genes were not required to have neuroanatomic abnormalities. Additionally, *Sap130* mutations presented with a more severe brain phenotype compared to *Pcdha9* mutations. This is interesting as knock-out mice for *Sap130* are embryonically lethal whereas *Pcdha9* knock-out mice are viable suggesting that *Sap130* may have a more upstream effect whereas *Pcdha9* is more downstream.^105^ *Sap130* functions as a subunit of the histone deacetylase dependent co-repressor complex which has roles transcriptional regulation and has been specifically mentioned to be required for recovery from hypoxia.^106^ *Pcdha9* is a member of the protocadherin alpha gene cluster which are important plasma membrane proteins for cell-cell connections, neural projects, and synapse formation.^107^ Interestingly both genes show overlap in connectomes with chromatin and histone modification pathways.^44^ Alterations in histone modifying pathways have been linked to various forms of CHD. ^108–110^ It’s likely that there are many other genes and gene pairings that could cause a similar neurodevelopmental phenotype as it’s possible the main driver could be epigenetic dysregulation resulting in altered differential expression of genes required for signaling and migration. Incorrect timing for migrating neural cells might explain why preterm babies and CHD babies look phenotypically similar in regard to neurodevelopment. Looking at the genetics of the preclinical mouse model could not be conclusive as to the etiology of the human CHD neurodevelopmental issues; however, we demonstrate spectra of cardiac and brain anatomical abnormalities associated with the different genetic combinations of the *Ohia* mouse, parallel to those seen in human subjects.

CHD subjects have a documented risk of poor neurodevelopmental outcomes although the specific mechanism is unknown.^111–115^ Additionally, neurodevelopmental impairments can continue to affect individuals well past infancy and into adolescence and early adulthood.^116–121^ There are two main hypotheses with regard to the origin of injury to the brain and subsequent neurodevelopmental deficits that arise from the injury; intrinsic patient factors including genetics or altered substrate delivery due to the cardiac malformation. There have been studies looking at the heritability of CHD to determine the genetic causes which resulted in the finding that CHD most likely has a genetic component. CHD is particularly common in genetic syndromes caused by aneuploidy defects; ranging anywhere from 20% -100% of individuals with trisomies 13, 18, and 21 as well as and monosomy X^122^, though these syndromic cases account for a relatively low amount of the total CHD population. Additionally, rates of CHD are increased when there are abnormal chromosomal structural syndromes such as deletions, insertions, and copy number variants.^123^ Known de novo single nucleotide polymorphisms account for approximately 10% of CHD cases, roughly the same amount that can be attributed to environmental factors.^123,124^ However, the vast majority of CHD individuals have an unknown origin of the cardiac defect as well as subsequent neurodevelopmental abnormalities. To overcome this missing data, work has begun looking at previously uncharacterized de novo mutations in mice, singling out genes with human homologs and which have high expression in the heart. When looking at only the homologous genes, researchers found that between CHD and control individuals, there was no significant difference between rates of generic de novo mutations; however, when comparing genes with high cardiac expression, CHD subjects had much higher rates of mutations in protein-altering mutations.^125^ This recent work into the genetic origins of CHD has shown that even when the cause of the insult is unknown, the cause of CHD is likely genetic possibly through the alteration of genes which affect similar downstream pathways.

Whether the genetic insult is the main driver or whether the mutations leave the individual susceptible to other forms of injury is unknown. Our group has previously looked at how the developing subcortical paralimbic-related structures of the brain can be linked to energy metabolism and found that subcortical morphological differences were not likely linked to prematurity or white matter injury alone, but that there was a linkage between brain metabolism and subcortical structural morphometry.^58^ Stemming from these findings, we believe that subplate vulnerability in the prenatal and immediate post-natal period can be informative to understanding the subcortical dysmaturation seen in CHD infants. Within CHD individuals, it’s been shown that preoperatively they have volumetric abnormalities present in the cerebellum and brainstem^16^ and additionally that abnormal cerebellar growth has been linked with worse outcomes in expressive communication and the adaptive development quotient in CHD individuals.^12^ Furthermore, there is an established link between subcortical morphological abnormalities and white matter vulnerability which can lead to injury and eventual worse neurodevelopmental outcomes.^17,20^ With the mounting evidence of the importance of subcortical structures for proper brain development, and the subcortical structures’ relationship to proper cortical development, inclusion of subcortical structures into our scoring system is necessary to characterize the entire picture of brain dysplasia. With nearly half of CHD cases still without a known genetic origin, identifying subtle cases of neurodevelopmental abnormalities in CHD individuals can better help us understand the downstream effects of CHD on brain development, ultimately furthering knowledge of the interplay between CHD and neurodevelopmental outcomes. Importantly, early interventional strategies have shown promise in helping CHD individuals with potential neurocognitive difficulties^126^ and early identification of the most vulnerable through proper screening is necessary to identify and target therapies toward highest risk individuals.

### Limitations

Our study does have some important limitations. The preterm and term CHD cohorts that we studied were heterogenous related to heart lesion subtypes. During this study, only a limited number of socio-demographic factors were prospectively collected and therefore analysis using these factors was unable to be carried out and future work correlating BDS score with pediatric/adolescent neurocognitive outcomes in relation to socio-demographic factors is needed. Our multi-site approach with different scanner platforms and vendors can lead to interscanner variance, but our secondary analyses did not find a correlation between scanner vendor or field strength and brain dysplasia score. Future work employing retrospective machine learning techniques like the empirical Bayes technique (Combat) can be used to reduce interscanner variance. Importantly, we were not able to measure extra-axial CSF fluid in the ECM imaging of the mouse mutants, which is an important component of the BDS score. Complete removal of CSF and infiltration of paraffin could have altered CSF volumes or the ventricle shape. With this in mind, we did not feel comfortable labeling these brain regions as containing CSF and instead labeled them as extra-axial space.

### Contributors

Conceptualization: WTR, JVS, GG, VL,

Data Curation: WTR, JVS, GG, VL,

Manuscript Draft Preparation: WTR, JVS, GG, VL,

Manuscript Editing: WTR, JVS, GG, VL,

Statistical Analysis: VL, VS

Human Quantitative Scoring and Reads: SS, AP

Human Volumetric Segmentation: WTR,

Human Cardiac Lesion Characterization: JVS,

Mouse Quantitative Scoring: WTR, YW, AP

Mouse Volumetric Segmentation: RS,AP

Supervision: CL, AP

Verification of Underlying Data: WTR

## Data Availability

All data produced in the present study are available upon reasonable request to the authors.
Preclinical mouse data can be made available upon request to corresponding author. Clinical data cannot be made public due to privacy issues, but limited data can be made available upon special request to the corresponding author with justification for the data request.

## Acknowledgments

This work was supported by the Department of Defense (W81XWH-16-1-0613), the National Heart, Lung, and Blood Institute (R01 HL152740-1, R01 HL128818-05), and the National Heart, Lung and Blood Institute with National Institute on Aging (R01HL128818-05 S1). Southern California Clinical and Translational Sciences Institute (NCATS) through Grant UL1TR0001855. Its contents are solely the responsibility of the authors and do not necessarily represent the official views of the NIH. We also acknowledge Additional Ventures for support (AP, VR, RC). VR is supported by the Saban Research Institute, Additional Ventures Foundation and NIH-NHLBI K01HL153942. NT is supported by Children’s Hospital Los Angeles Clinical Services Research Grant and the NINR K23 Grant 1K23NR019121-01A1.

## Data Sharing Statement

Preclinical mouse data can be made available upon request to corresponding author. Clinical data cannot be made public due to privacy issues, but limited data can be made available upon special request to the corresponding author with justification for the data request.

## Non-standard Abbreviations and Acronyms

ECM: Episcopic Confocal Microscopy
CHD: congenital heart disease
HLHS: hypoplastic left heart syndrome
MRI: magnetic resonance imaging
WMI: white matter injury
GA: gestational age
PCA: post-conceptional age
BDS: brain dysplasia score.

## Supplemental Figure Legends

**Supplemental Figure 1.**
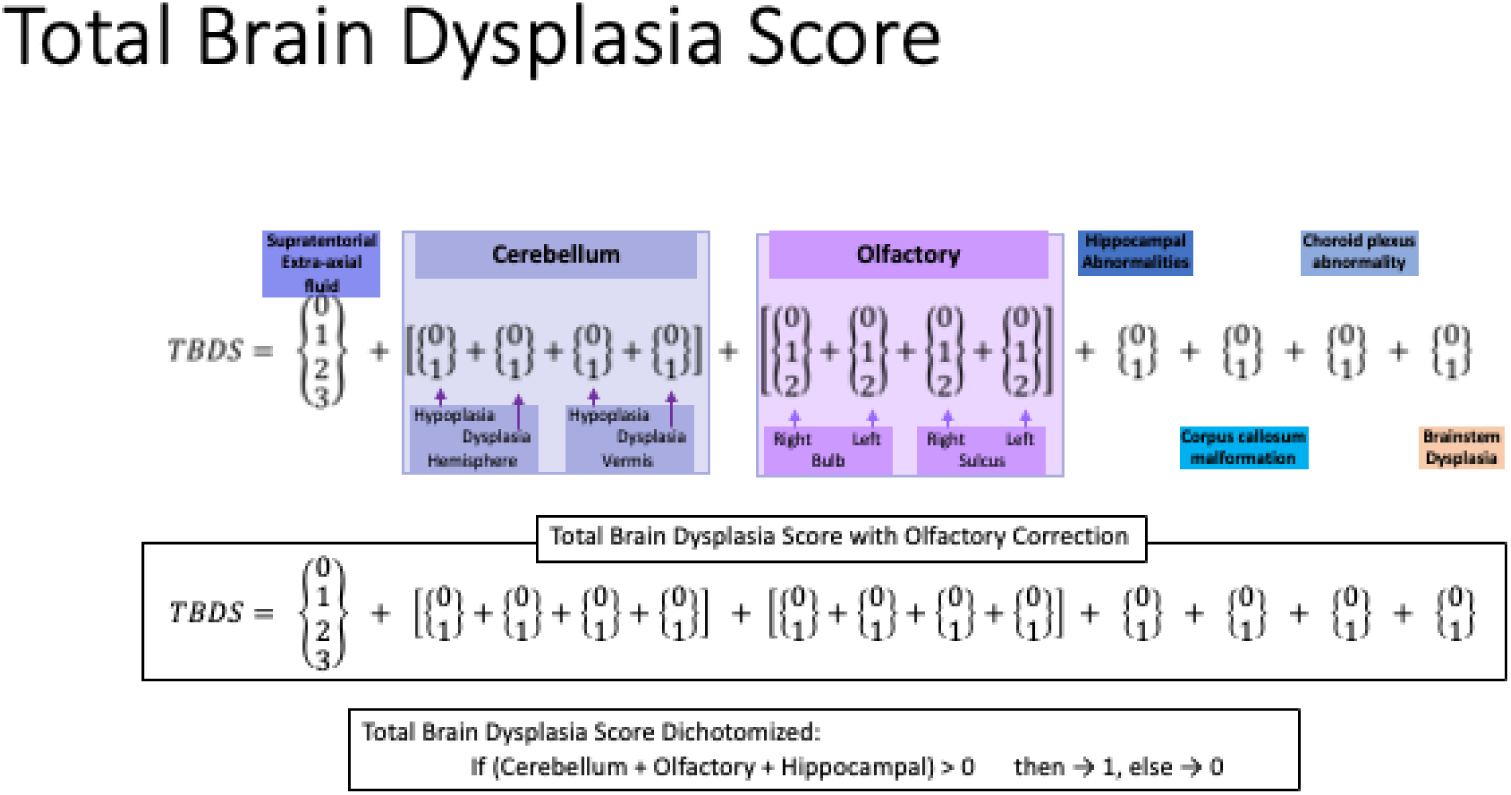
Components of Human Brain Dysplasia Scoring Criteria. Our brain dysplasia scoring criteria consisted of 13 distinct observations. Supratentorial extra-axial fluid was assessed for normality, cerebellar hemisphere and vermis were assessed for hypoplasia and dysplasia, right and left olfactory bulb and olfactory sulcus were examined for any abnormalities. Additionally, the hippocampus, corpus collosum, choroid plexus, and brainstem were checked for abnormalities.

**Supplemental Figure 2.**
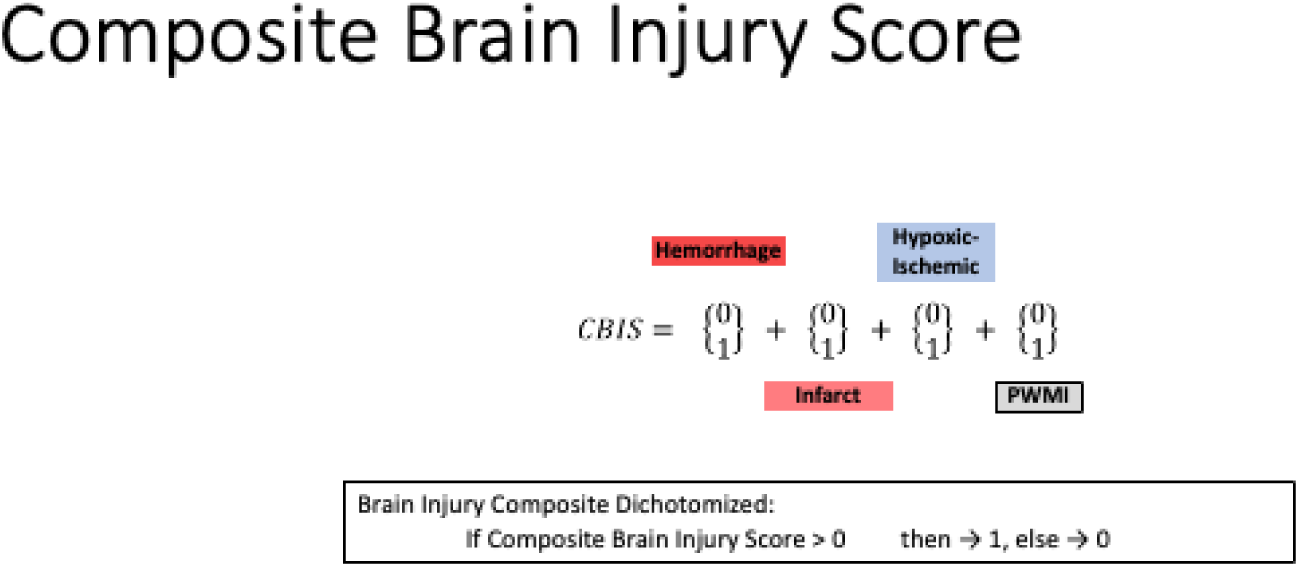
Components of Composite Brain Injury Score. Brain injury score was composed of four measures: hemorrhage, infarct, hypoxic ischemia, and perinatal white matter injury. Brain injury score was binary, if any injury was present in the above measures, the subject was considered to have a brain injury.

**Supplemental Figure 3.**
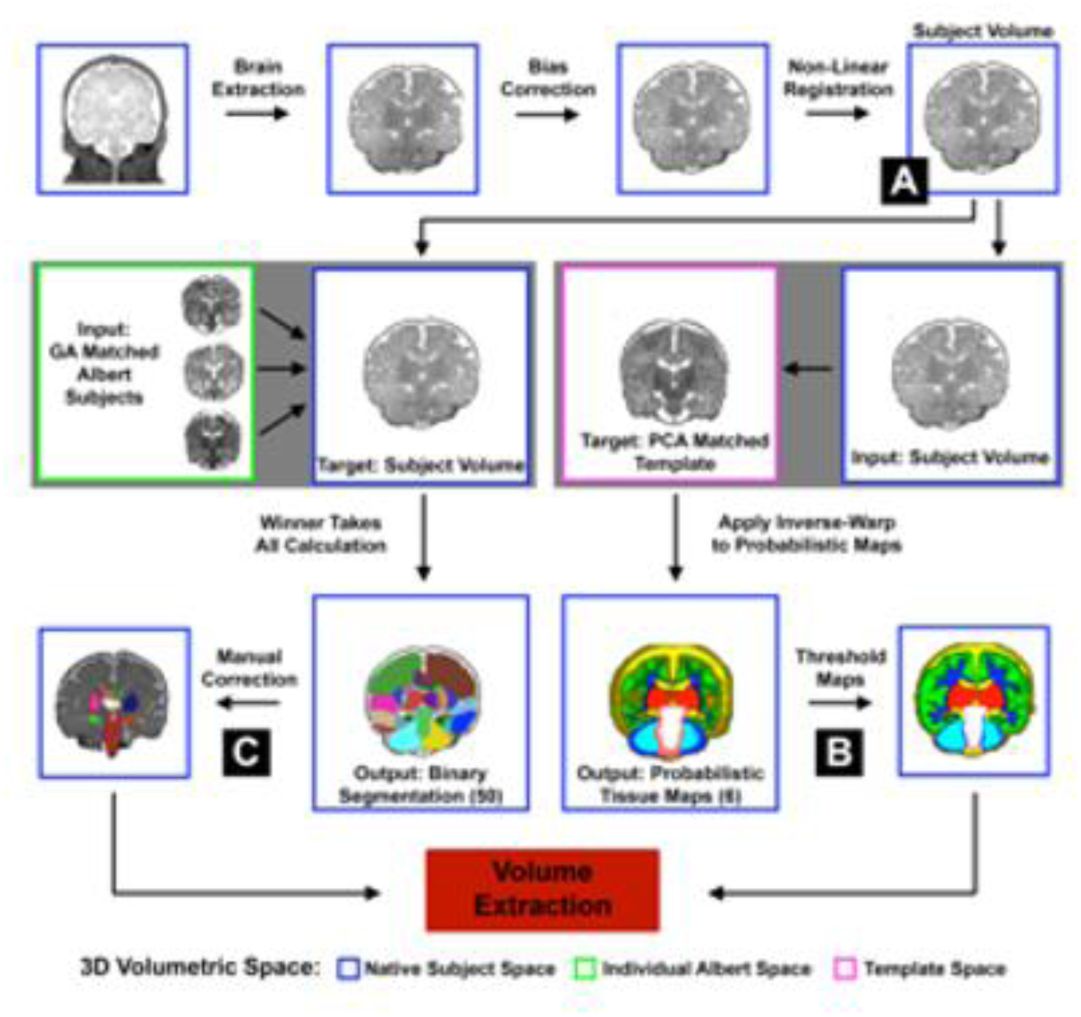
Flow Diagram for NeBSS, a Semi-automated Neonatal Segmentation Pipeline. Briefly, subject MR images are input into NeBSS which has two branches. Branch A uses the Albert Neonatal Atlas and outputs 50 distinct volumetric brain structures in the subject space. Branch B uses the Serag Neonatal Brain Atlas probability maps to output 10 volumetric brain regions.

**Supplemental Figure 4.**
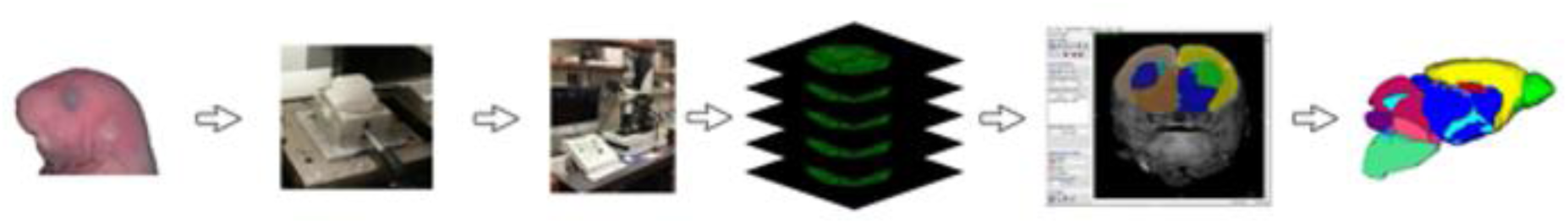
Flow Diagram of Sample Preparation, ECM Analysis, and Volumetric Analysis. Samples selected for analysis were first necropsied. Post-necropsy samples were dehydrated using a Sakura Tissue Tek system and processed on the ECM machine. Samples were then manually segmented using ITK-Snap before volumetric results were analyzed.

**Supplemental Figure 5.**
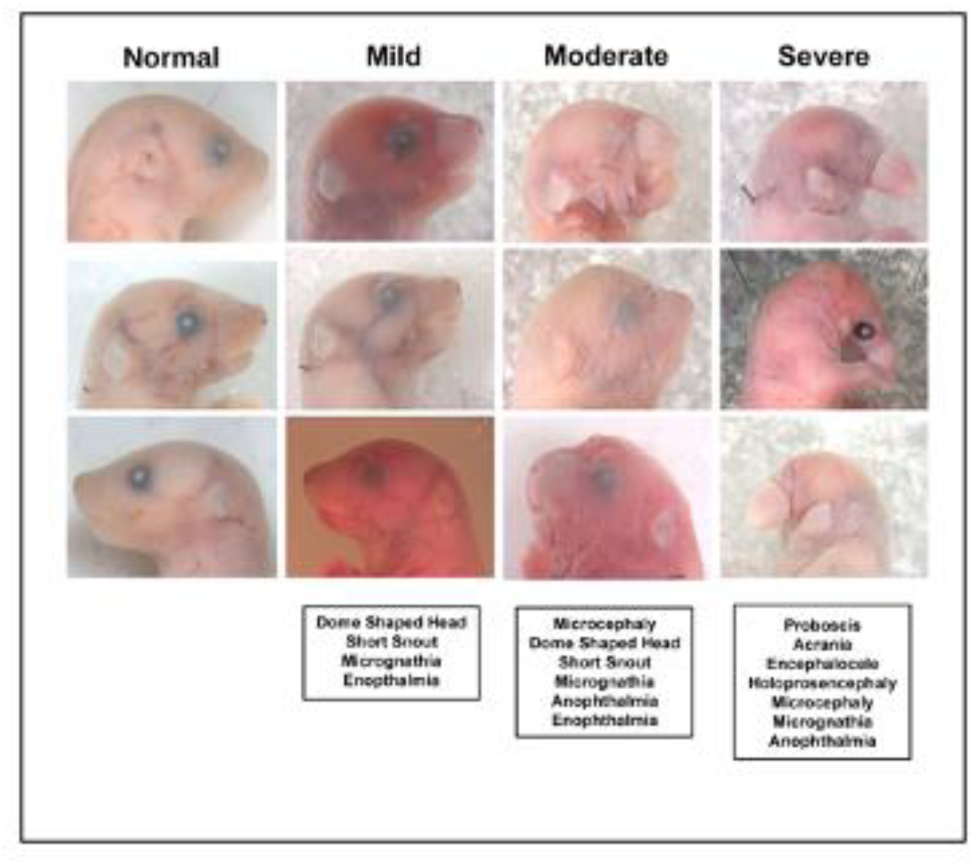
Necropsy Among Mutant Mice Showed Variety and Differing Degree of Severity of Malformations. Mice in the mouse screen presented with craniofacial anomalies with varying degrees of severity. More mild forms of errant craniofacial anatomy presented with mild defects in the ears and eyes. Moderate forms presented as more severe ear and eye defects as well as a shortened snout. Lastly, severe defects presented with large structural defects in the craniofacial anatomy, often with underlying structural brain defects as well.

**Supplemental Figure 6.**
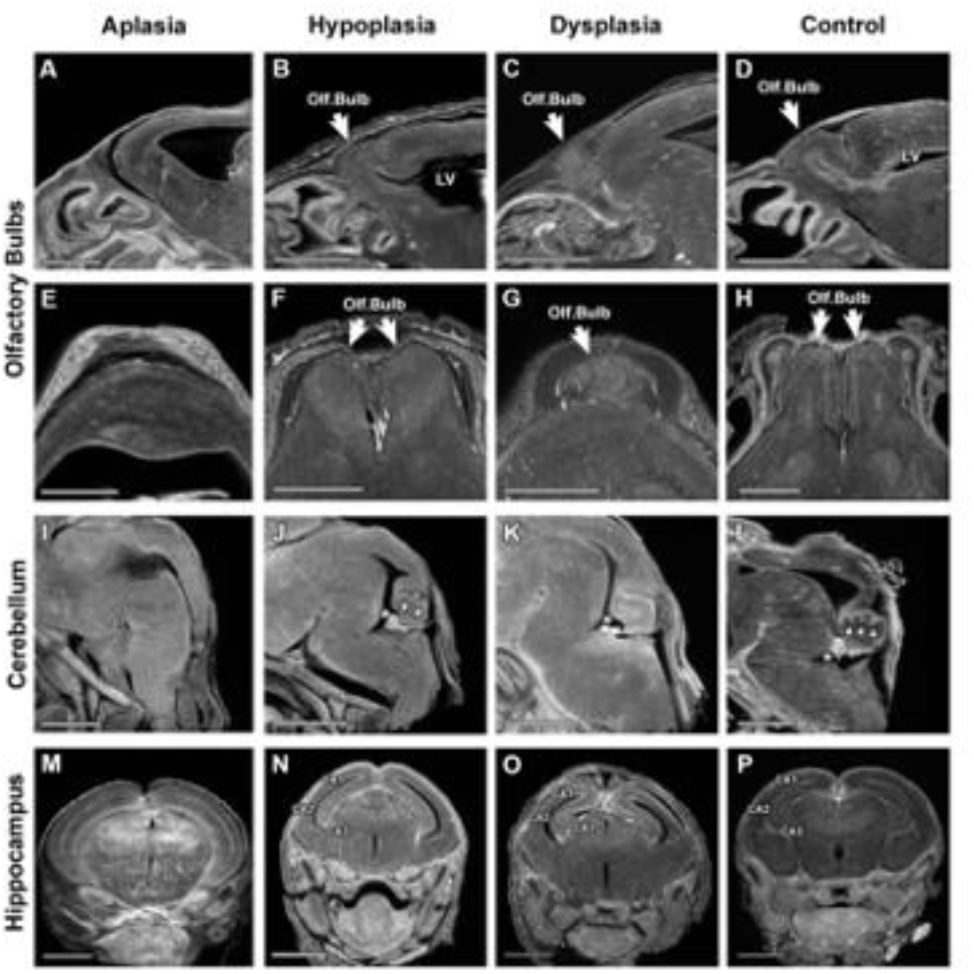
Individual Structures Showed a Range of Abnormality in Screen Mice. Episcopic Fluorescence Image Capture (ECM) was carried out on prepared mouse brains. Samples presented with varying degrees of abnormalities. Three major structures analyzed were olfactory bulbs, cerebellum, and hippocampus. Within each structure, mice had a variety of defects including aplasia, hypoplasia, and dysplasia.

**Supplemental Figure 7.**
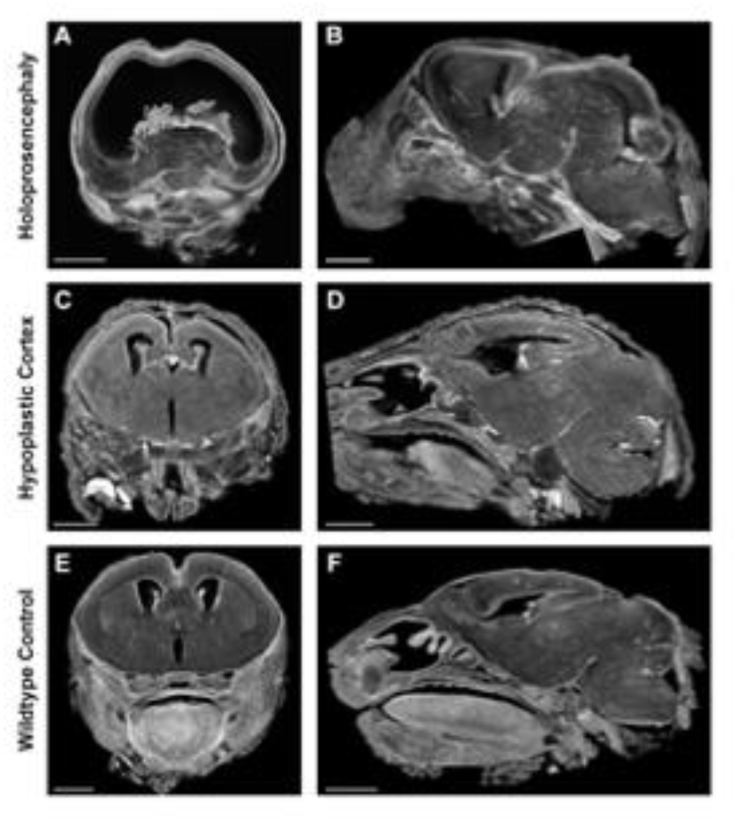
Varying Degree of Cerebral Abnormalities Present in Screen Mice. ECM analysis found that mice had varying degrees of severity of cerebral abnormalities including holoprosencephaly and cerebral hypoplasia.

**Supplemental Figure 8.**
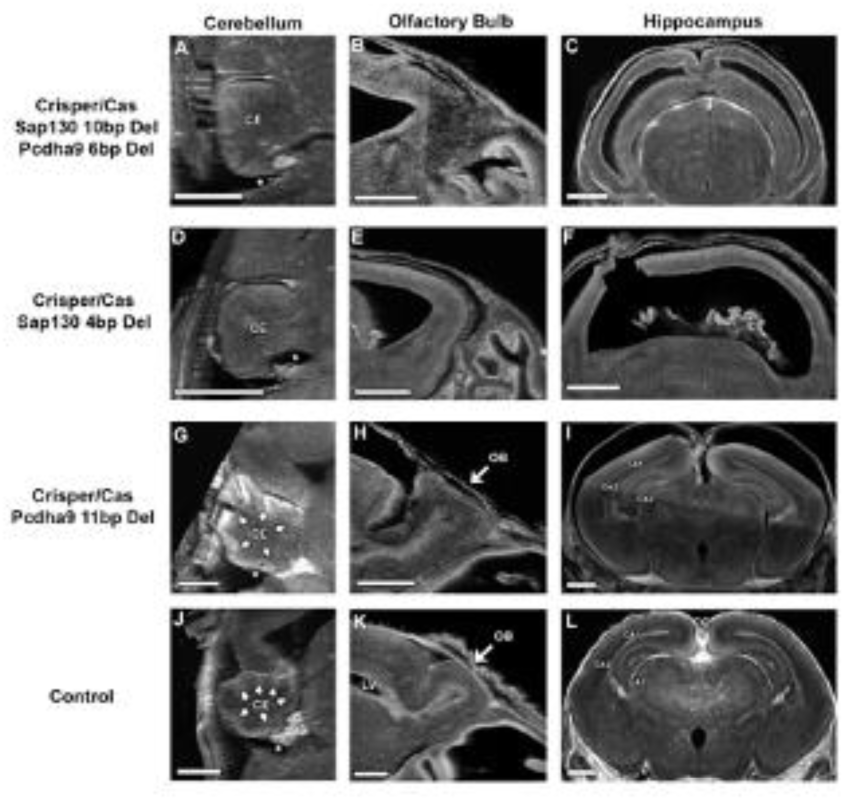
Crispr/Cas Figure Mutant Animals Showed Similar Phenotype to Screen Mice. Using ECM, Crispr/Cas9 mutants were analyzed and show similar phenotypes seen in mutant mice recovered from the phenotype screen. A similar pattern of dysplasia and aplasia were seen in the cerebellum, olfactory bulb and hippocampus of mice recovered in the screen.

**Supplemental Figure 9.**
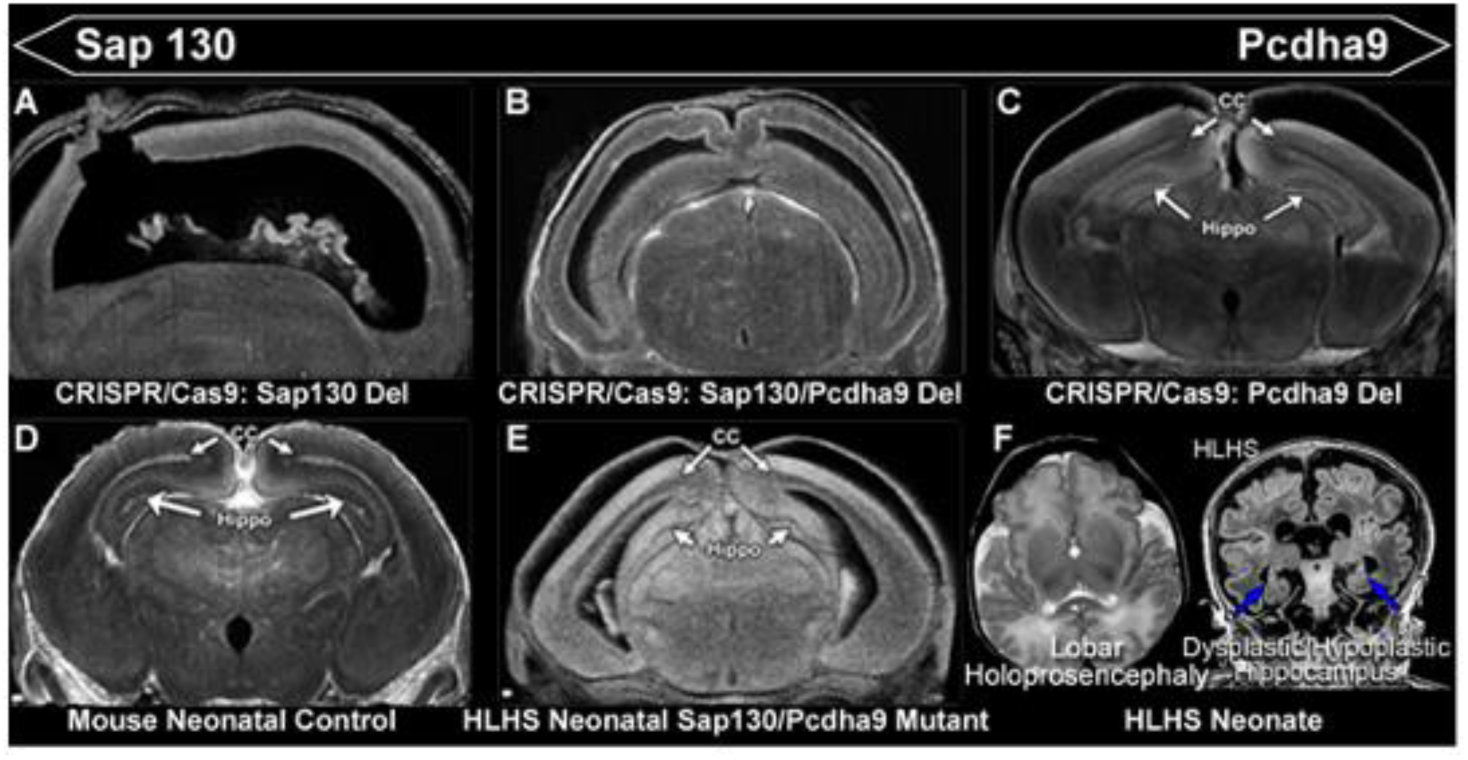
Holoprosencephaly and Hippocampal Malformations Seen in Human and Mouse. A, Alobar holoprosencephaly seen in Sap130 CRISPR/Cas9 deletion mouse. B, Lobar holoprosencephaly seen in Sap130/Pcdha9 double CRISPR/Cas9 deletion mouse. C, Pcdha9 CRISPR/Cas9 deletion mouse displaying cerebral hypoplasia and dysmaturation as well as hippocampal abnormalities. D, Control mouse showing normal cortical and hippocampal anatomy. E, Sap130/Pcdha9 double mutant mouse exhibiting cortical, corpus collosum, and hippocampal abnormalities. F, Lobar holoprosencephaly and hippocampal malformations observed in human subjects with HLHS.

