## Supplemental Tables for "Development and Validation of a Paralimbic Related Subcortical Brain Dysmaturation MRI Score in Infants with Congenital Heart Disease"

**Supplemental Table 1. Incidence of Brain Injury in Human Infant Term and Preterm CHD**

| Injury (Dichotomous) | Preterm CHD (n=69) | Preterm Non-CHD (n=51) | Comparison | Term CHD (n =265) | Term Controls (n=93) | Comparison |
| --- | --- | --- | --- | --- | --- | --- |
|  | **Number of Subjects with Injury (%)** | | **p-value** | **Number of Subjects with Injury (%)** | | **p-value** |
| Hemorrhage | 10 (14.71%) | 9 (17.65%) | 0.6647 | 19 (7.20%) | 1 (1.09%) | **0.0284** |
| Focal Infarct | 6 (8.82%) | 0 (0.00%) | **0.0295** | 23 (8.68%) | 0 (0.00%) | **0.0035** |
| Hypoxic Ischemic Injury | 2 (3.03%) | 0 (0.00%) | 0.2144 | 8 (3.15%) | 0 (0.00%) | 0.1038 |
| Punctate White Matter Lesions | 12 (17.39%) | 2 (3.92%) | **0.0231** | 40 (15.09%) | 0 (0.00%) | **<0.0001** |
| *Dichotomized Injury Composite* | 25 (36.23%) | 11 (21.57%) | 0.0831 | 74 (27.92%) | 1 (1.08%) | **<0.0001** |
| Injury (Categorical) | **Mean *(Standard Error)*** | | **p-value** | **Mean *(Standard Error)*** | | **p-value** |
| *Injury Composite* | 0.434783 *(0.075797)* | 0.215686 *(0.058166)* | 0.0581 | 0.339623 *(0.037319)* | 0.010753 *(0.010753)* | **<0.0001** |

**Supplemental Table 2: Comparison of Cortical Maturation Score (TMS) between CHD and Controls Infants (Preterm and Term)**

| BDS Maturation (Categorical) | Preterm CHD (*n* = 69) | Preterm Non-CHD (*n* = 51) | Comparison | Term CHD (*n* = 260) | Term Control (*n* = 93) | | Comparison |
| --- | --- | --- | --- | --- | --- | --- | --- |
|  | **Mean *(Standard Error)*** | | **p-value** | **Mean *(Standard Error)*** | | | **p-value** |
| Occipital Cortex: Cortical Folding | 3.0435 *(0.1041)* | 2.8627 *(0.1284)* | 0.3095 | 2.9577 *(0.0515)* | | 2.7849 *(0.1473)* | 0.6235 |
| Frontal Cortex: Cortical Folding | 2.5942 *(0.1359)* | 2.4902 *(0.1735)* | 0.6604 | 1.2366 *(0.0786)* | | 2.6731 *(0.0710)* | **<0.0001** |
| Insular Cortex: Cortical Folding | 2.9855 *(0.1195)* | 3.0980 *(0.1407)* | 0.5044 | 3.1538 *(0.0537)* | | 4.3441 *(0.0692)* | **<0.0001** |
| Frontal Cortex: Dark Bands on T2 | 1.8986 *(0.1477)* | 1.9020 *(0.1820)* | 0.8021 | 2.2335 *(0.0738)* | | 3.6882 *(0.0865)* | **<0.0001** |
| Parietal Cortex: Dark Bands on T2 | 2.2899 *(0.1587)* | 2.6863 *(0.2029)* | 0.1345 | 2.8308 *(0.0746)* | | 3.9140 *(0.0423)* | **<0.0001** |
| Myelination: Dark Bands on T2 | 3.5735 *(0.1297)* | 4.1176 *(0.1143)* | **0.0049** | 4.0888 *(0.0603)* | | 5.4409 *(0.0725)* | **<0.0001** |

**Supplemental Table 3: Inter-rater Reliability (Kappa) for Brain Dysplasia Score Between Two Pediatric Neuroradiologists**

|  | κ (kappa) Reviewer 1 | κ (kappa) Reviewer 2 |
| --- | --- | --- |
| Microcephaly Yes/No | 1 | 0.4167 |
| Macrocephaly Yes/No | 1 | 1 |
| Cerebellar R/L Hemispheric Hypoplasia | 1 | 0.6786 |
| Cerebellar R/L Hemispheric Dysplasia | 1 | 0 |
| Cerebellar Vermian Hypoplasia Yes/No | 0.708333 | 0.6434 |
| Cerebellar Vermian Dysplasia Yes/No | 1 | 0.2895 |
| Supratentorial Extra-axial fluid | 0.350962 | 0.0526 |
| Maturational Parameters: Cortical Folding | 0.18306 | 0 |
| Maturational Parameters: Frontal | 0.371257 | 0.3698 |
| Maturational Parameters: Insular Cortex | 0.310627 | 0.3413 |
| Dark bands on T2 (cellular): Frontal | 0.638365 | 0.3061 |
| Dark bands on T2 (cellular): parietal | 0.904959 | 0.2140 |
| Dark bands on T2 (cellular): Myelination [M] | 0.350427 | 0.0256 |
| Dark bands on T2 (cellular): Germinal Matrix | 0.480226 | 0 |
| Dysmorphometry: Right Olfactory Bulb | 0.893773 | 0.6346 |
| Left Olfactory Bulb | 0.843243 | 0.7373 |
| Right Olfactory Sulci | 1 | 0.6329 |
| Left Olfactory Sulci | 1 | 0.6949 |
| Micrognathia | 0.68272 | 0.4408 |
| Midface hypoplasia | 1 | 1 |
| Temporal bone abnormalities | 1 | 0.5385 |
| Orbital abnormality | 0.649351 | 0.4000 |
| Hippocampal abnormalities | 0.910217 | 0.5190 |
| Cortical thickness abnormalities | 1 | 0.4633 |
| Corpus callosum malformation | 0.888031 | 0.5957 |
| Corpus callosum volume abnormalities | 0.828402 | 0.3284 |
| Ventriculomegaly | 0.538462 | 0.3640 |
| Absent Septum pellucidum | 0.606335 | 0.9474 |
| Choroid plexus abnormality | 0.125628 | 0.4311 |
| Brainstem Dysplasia | 0.888031 | 0.5342 |
| Holoprosencephaly3 | 1 | 1 |
| Interdigitation of medial gyri | 1 | 1 |
| Injury: Hemorrhage | 0.867925 | 0.7692 |
| Infarct | 1 | 1 |
| Hypoxic-ischemic central BG/thal/perirolandic injury pattern | 0.648649 | 1 |
| Puncate white matter lesion Yes/No | 0.626667 | 0.7805 |

**Supplemental Table 4A: Association Between Innate Factors, Heart Lesion Subtypes and Brain Dysplasia Score in Term CHD**

| Clinical Characteristics (Total Composite Correlates) | *R^2^* | p-value *(estimate)* |
| --- | --- | --- |
| Birth Weight (g) | 0.0007 | 0.8186 *(-0.0001)* |
| Birth Weight Percentile | 0.0110 | 0.3538 *(-0.0107)* |
| Head circumference (cm) | 0.0396 | 0.0785 *(0.1712)* |
| Head circumference Percentile | 0.0302 | 0.1255 *(0.0175)* |
| Birth Length (cm) | 0.0137 | 0.3140 *(0.0520)* |
| Birth Length Percentile | 0.0001 | 0.9386 *(-0.0009)* |
| APGAR (1 minute) | 0.0001 | 0.9417 *(-0.0142)* |
| APGAR (5 minutes) | 0.0074 | 0.4670 *(0.1899)* |
| 22q11 microdeletion | 0.0294 | 0.1862 *(-1.7758)* |
| Single Ventricle | 0.0052 | 0.5245 *(-0.4444)* |
| Aortic arch Obstruction | 0.0055 | 0.5118 *(0.4616)* |
| Double Ventricle with Arch Obstruction | 0.0091 | 0.4009 *(0.9202)* |
| Single Ventricle with Arch Obstruction | 0.0018 | 0.7118 *(0.2764)* |
| d-Transposition of Great Arteries | 0.0026 | 0.6547 *(0.3345)* |
| Conotruncal cardiac defect | 0.0104 | 0.3683 *(0.6354)* |
| Altered fetal cerebral substrate delivery | 0.0092 | 0.4069 *(0.9420)* |
| Altered fetal cerebral substrate delivery, severity score | 0.0143 | 0.3114 *(1.2857)* |
| Heterotaxy | 0.0219 | 0.1903 *(1.8667)* |

**Supplemental Table 4B:Association Between Preoperative Risk Factors and Brain Dysplasia Score**

| Preoperative Factors (Total Composite Correlates) | *N with data* | *R^2^* | p-value *(estimate)* |
| --- | --- | --- | --- |
| Preoperative Arterial Blood Gas (ABG) pH | 53 | 0.0316 | 0.2029 *(-8.3331)* |
| Preoperative ABG PaO2 | 51 | 0.0053 | 0.6130 *(-0.0106)* |
| Preoperative ABG Lactate (mmol/L) | 63 | 0.0015 | 0.7598 *(0.1539)* |
| Preoperative Renal Dysfunction (CR > 1) | 75 | 0.0000 | 0.9661 *(0.1351)* |
| Preoperative Hepatic Dysfunction (INR > 2) | 65 | 0.0092 | 0.4485 *(-1.7460)* |
| Preoperative Inotrope Use | 80 | 0.0008 | 0.8043 *(-0.1764)* |
| Age at Surgery (postnatal days) | 78 | 0.0001 | 0.9393 *(-0.0009)* |
| Age at Surgery ≤7 days | 78 | **0.0503** | **0.0484 *(-1.4231)*** |
| Age at Surgery (post conceptual age – weeks) | 78 | 0.6598 | 0.4347 *(-3.0000)* |

**Supplemental Table 4C Human Infant Association Between Intraoperative Factors and Brain Dysplasia Score**

| Intraoperative Factors (Total Composite Correlates) | *N with data* | *R^2^* | p-value *(estimate)* |
| --- | --- | --- | --- |
| Bypass Procedure | 90 | 0.0044 | 0.5348 *(0.6421)* |
| Bypass Time (minutes) | 82 | 0.0070 | 0.4563 *(0.0046)* |
| Aortic Cross-Clamp Procedure | 86 | 0.0087 | 0.3932 *(0.6134)* |
| Aortic Cross-Clamp Time (minutes) | 65 | 0.0094 | 0.4430 *(0.0075)* |
| Circulatory Arrest / DHCA Procedure | 86 | 0.0037 | 0.5802 *(0.3885)* |
| Circ arrest / DHCA Time (minutes) | 73 | 0.0001 | 0.9205 *(0.0019)* |

**Supplemental Table 4D. Human Association Between Post-Operative Clinical Risk Factors and Brain Dysplasia Score**

| Clinical Risk Factors (Total Composite Correlates) | N with data | R^2^ | p-value *(estimate)* |
| --- | --- | --- | --- |
| ECMO 1st hospitalization | 87 | 0.0365 | 0.0764 *(-1.6194)* |
| ECMO (days) | 62 | 0.0251 | 0.2190 *(0.3209)* |
| Delayed Sternal Closure | 90 | 0.0006 | 0.8263 *(0.1662)* |
| Number of Cardiac Surgeries Lifetime | 90 | 0.0033 | 0.5908 *(-0.2136)* |
| Unplanned interventions 1st hospitalization | 88 | 0.0023 | 0.6597 *(0.3063)* |
| Length of ICU stay (days) | 90 | 0.0373 | 0.0681 *(0.0190)* |
| Length of Hospitalization (days) | 90 | **0.0447** | **0.0454 *(0.0180)*** |
| Expired 1st hospitalization | 90 | 0.0224 | 0.1596 *(-2.3023)* |
| CPR 1st hospitalization | 90 | 0.0021 | 0.6678 *(0.6353)* |
| Seizures in ICU | 90 | 0.0090 | 0.3748 *(-0.8289)* |
| Home anti-epileptics | 90 | 0.0143 | 0.2610 *(-1.1600)* |
| Home with G-tube | 90 | **0.0611** | **0.0189 *(-1.9214)*** |
| Home with tracheostomy / ventilator | 90 | 0.0176 | 0.2131 *(-2.0407)* |

**Supplemental Table 5: Association Between Field Strength and Brain Dysplasia Score**

| Brain Dysplasia Score:  Abnormalities / Injury / Maturation | Satterthwaite | | ANOVA | | Satterthwaite | | ANOVA | |
| --- | --- | --- | --- | --- | --- | --- | --- | --- |
|  | **Preterm**  **CHD & Control p-value** | **Term**  **CHD & Control p-value** | **Preterm**  **CHD & Control p-value** | **Term**  **CHD & Control p-value** | **Preterm**  **CHD p-value** | **Term**  **CHD p-value** | **Preterm**  **CHD p-value** | **Term**  **CHD p-value** |
| Bilateral Cerebellar Hemispheric Hypoplasia | 0.0992 | 0.6417 | 0.5888 | **0.0126** | **0.0495** | 0.6832 | 0.2660 | 0.1800 |
| Bilateral Cerebellar Hemispheric Dysplasia | 0.0901 | **0.0004** | 0.2167 | **0.0002** | **0.0123** | **0.0005** | 0.1048 | **0.0001** |
| Cerebellar Vermis Hypoplasia | **0.0370** | 0.0846 | 0.6773 | **0.0019** | **0.0192** | 0.6006 | 0.4229 | 0.1263 |
| Cerebellar Vermis Dysplasia | **0.0033** | **0.0001** | **0.0245** | **0.0007** | **0.0033** | **0.0003** | 0.0601 | **0.0003** |
| *Cerebellum Composite* | 0.8107 | **0.0008** | 0.4113 | **0.0015** | 0.5311 | **0.0120** | 0.7134 | **0.0109** |
| *Dichotomized Cerebellum Composite* | 0.6801 | **0.0021** | 0.3437 | **0.0011** | 0.3936 | 0.0664 | 0.6636 | **0.0389** |
| Right Olfactory Bulb | 0.9840 | **0.0055** | 0.7455 | **<0.0001** | 0.4382 | 0.3406 | 0.4817 | **0.0435** |
| Left Olfactory Bulb | 0.8473 | **0.0055** | 0.6288 | **<0.0001** | 0.5612 | 0.3012 | 0.5761 | **0.0252** |
| Right Olfactory Sulcus | 0.9673 | **0.0007** | 0.6053 | **0.0023** | 0.5073 | 0.1067 | 0.6688 | 0.1043 |
| Left Olfactory Sulcus | 0.9115 | **0.0005** | 0.5670 | **0.0012** | 0.5513 | 0.1067 | 0.7082 | 0.1043 |
| *Olfactory Composite* | 0.7447 | **0.0011** | 0.8922 | **0.0001** | 0.3158 | 0.0928 | 0.2464 | **0.0221** |
| *Dichotomized Olfactory Composite* | 0.8473 | **0.0061** | 0.6288 | **<0.0001** | 0.5612 | 0.3215 | 0.5761 | **0.0348** |
| Hippocampus | 0.5216 | **0.0013** | 0.6854 | **<0.0001** | 0.2794 | 0.2063 | 0.9440 | **0.0125** |
| Corpus Callosum | **0.0398** | **0.0048** | **0.0172** | **0.0194** | 0.0666 | **0.0192** | **0.0428** | **0.0063** |
| Choroid Plexus | 0.8474 | **0.0070** | 0.4794 | 0.6707 | 0.9468 | 0.1532 | 0.5233 | 0.6467 |
| Brainstem | 0.3111 | **<0.0001** | 0.3583 | **<0.0001** | 0.5286 | **<0.0001** | 0.6414 | **<0.0001** |
| Supratentorial Extra-Axial Fluid | **0.0016** | **0.0157** | **0.0383** | **0.0001** | **0.0012** | 0.3355 | **0.0225** | **0.0327** |
| Hemorrhage | 0.3347 | 0.8587 | 0.5106 | 0.7653 | 0.4820 | 0.3420 | 0.7880 | 0.5194 |
| Infarct | 0.5397 | 0.1225 | 1.0000 | **0.0271** | 0.7489 | 0.3461 | 0.6952 | **0.0331** |
| Hypoxic Ischemic Injury | 0.8730 | 0.1968 | 0.5849 | 0.1611 | 0.7639 | 0.3109 | 0.4337 | 0.0926 |
| Punctate White Matter Lesions (PWM) | 0.8257 | **0.0266** | 0.1972 | 0.3759 | 0.8279 | 0.1580 | 0.4210 | 0.4350 |
| PWM: Laterality | 0.5384 | 0.8623 | 0.7114 | 0.7785 | 0.5384 | 0.8623 | 0.7114 | 0.7785 |
| PWM: Distribution | 0.2889 | 0.3115 | 0.1885 | 0.8495 | 0.2889 | 0.3115 | 0.1885 | 0.8495 |
| PWM: Lobes | 0.9155 | 0.1419 | 0.1588 | 0.1552 | 0.9155 | 0.1419 | 0.1588 | 0.1552 |
| PWM: Spatially Co-incident with Banding | . | **0.0261** | 0.6887 | **<0.0001** | . | 0.3678 | 0.6887 | **0.0330** |
| *Injury Composite* | 0.7851 | **0.0114** | 0.8366 | 0.1274 | 0.5979 | 0.2012 | 0.9403 | 0.1086 |
| *Dichotomized Injury Composite* | 0.6479 | **0.0088** | 0.5743 | 0.2336 | 0.5135 | 0.2551 | 0.4066 | 0.0533 |
| Occipital Cortex: Cortical Folding | **0.0341** | 0.1383 | 0.5319 | **0.0016** | 0.0531 | 0.1043 | 0.8062 | 0.0986 |
| Frontal Cortex: Cortical Folding | **0.0117** | **0.0256** | 0.0558 | **0.0003** | **0.0106** | 0.6073 | 0.0611 | 0.5007 |
| Insular Cortex: Cortical Folding | 0.7842 | **0.0055** | 0.3428 | **<0.0001** | 0.4037 | 1.0000 | 0.7304 | 0.0507 |
| Frontal Cortex: Dark Bands on T2 | 0.0828 | **0.0466** | **0.0191** | **<0.0001** | 0.1625 | 0.5799 | **0.0477** | 0.3573 |
| Parietal Cortex: Dark Bands on T2 | 0.6331 | 0.8653 | 0.2648 | **<0.0001** | 0.7610 | **0.0352** | 0.3236 | **0.0052** |
| Myelination: Dark Bands on T2 | 0.8579 | **0.0001** | 0.3030 | **0.0357** | 0.3318 | 0.4581 | 0.7365 | 0.2596 |
| Germinal Matrix: Dark Bands on T2 | 0.1045 | 0.6032 | 0.2885 | 0.2352 | **0.0326** | 0.1555 | 0.0537 | **0.0474** |
| *Total Composite* | 0.5501 | **<0.0001** | 0.7548 | **<0.0001** | 0.1787 | **0.0127** | 0.5573 | **0.0069** |

** Welch–Satterthwaite t-test was used to test volume differences among individual structures.*

**Supplemental Table 7: Ohia Mouse Mutant Cohort Regional Cerebral Volumes Compared to Wild Type (WT)**

|  | | | **Raw (Non-normalized) Volume** | | | **Normalized Volume** | | |
| --- | --- | --- | --- | --- | --- | --- | --- | --- |
| **Structure** | N Ohia | N WT | Ohia Mean (x10^8^ μm^3^) | WT Mean (x10^8^ μm^3^) | P value * | Ohia Percent TBV | WT Percent TBV | P value * |
| **Total Volume** | 25 | 10 | 45.88 | 54.5 | 0.297 | . | . | . |
| **Intraventricular Vol.** | 11 | 9 | **1.968** | 0.531 | **0.009** | **0.043** | 0.011 | **0.015** |
| **Supratentorial** | 25 | 10 | 34.68 | **68.46** | **0.037** | 0.745 | **1.287** | **0.01** |
| **Infratentorial** | 25 | 10 | 10 | 11.81 | 0.358 | 0.232 | 0.212 | 0.208 |
| **L. Hippocampus** | 21 | 10 | 0.852 | 1.039 | 0.336 | 0.017 | 0.019 | 0.293 |
| **R. Hippocampus** | 20 | 10 | 0.831 | 0.981 | 0.431 | 0.016 | 0.018 | 0.406 |
| **L. Olfactory Bulb** | 18 | 10 | 2.017 | 2.212 | 0.59 | 0.038 | 0.041 | 0.121 |
| **R. Olfactory Bulb** | 17 | 10 | 2.066 | 2.188 | 0.711 | 0.038 | 0.041 | 0.057 |
| **L. Subcortical** | 25 | 10 | 4.104 | **5.884** | **0.041** | 0.097 | 0.107 | 0.223 |
| **R. Subcortical** | 25 | 10 | 4.073 | **5.805** | **0.044** | 0.096 | 0.106 | 0.178 |
| **L. Cortex** | 24 | 10 | 8.159 | 9.133 | 0.514 | 0.167 | 0.171 | 0.725 |
| **R. Cortex** | 23 | 10 | 8.192 | 9.392 | 0.445 | 0.162 | 0.174 | 0.295 |
| **Cerebellum** | 24 | 9 | 1.624 | **2.451** | **0.005** | 0.038 | 0.043 | 0.095 |
| **Pons** | 25 | 10 | 2.205 | 2.656 | 0.37 | 0.052 | 0.048 | 0.367 |
| **Medulla** | 24 | 10 | 6.459 | 6.803 | 0.77 | **0.15** | 0.122 | **0.024** |
| **Hypothalamus** | 7 | 10 | 0.118 | 0.148 | 0.267 | 0.002 | 0.003 | 0.074 |
| **Choroid Plexus** | 24 | 10 | 0.232 | 0.228 | 0.959 | 0.006 | 0.004 | 0.189 |
| **Midbrain** | 25 | 10 | 5.802 | 5.346 | 0.605 | **0.138** | 0.098 | **0.005** |

**Supplemental Table 6A: Brain Dysplasia Score within Ohia Mouse Mutant Cohort: Comparison between Cardiac Lesion Subgroups**

|  | CHD vs. No CHD | | | | Single vs. Biventricular Morphology | | | |
| --- | --- | --- | --- | --- | --- | --- | --- | --- |
| Structure | N | CHD | No CHD | P value * | N | Single | Double | P value * |
| Aplastic Hippocampus | 68 | 8 | 0 | 0.050 | 49 | 0 | 8 | 0.534 |
| Hypoplastic Hippocampus | 68 | 10 | 4 | 0.868 | 48 | 1 | 9 | 0.310 |
| Combination Hippocampus | 69 | 43 | 9 | 0.059 | 49 | 2 | 41 | 0.955 |
| Dysplastic Hippocampus | 68 | **33** | 9 | **0.043** | 48 | 2 | 31 | 0.341 |
| Hypoplastic Cerebrum | 68 | **13** | 0 | **0.008** | 48 | 0 | 13 | 0.389 |
| Dysplastic Cerebrum | 68 | **35** | 6 | **<0.001** | 48 | 2 | 33 | 0.489 |
| Aplastic Cerebellum | 69 | 1 | 1 | 0.538 | 49 | 1 | 0 | **<0.001** |
| Hypoplastic Cerebellum | 67 | 6 | 2 | 0.707 | 47 | 1 | 5 | 0.112 |
| Combination Cerebellum | 69 | 42 | 9 | 0.818 | 49 | 2 | **40** | **<0.001** |
| Dysplastic Cerebellum | 67 | **40** | 7 | **<0.001** | 47 | 1 | 39 | 0.161 |
| Aplastic L. Olf. Bulb | 69 | **30** | 2 | **<0.001** | 49 | 2 | 28 | 0.260 |
| Hypoplastic L. Olf. Bulb | 67 | 7 | 2 | 0.553 | 47 | 0 | 7 | 0.556 |
| Combination L. Olf. Bulb | 69 | **37** | 4 | **<0.001** | 49 | 2 | 35 | 0.292 |
| Dysplastic L. Olf. Bulb | 67 | 2 | 0 | 0.345 | 47 | 0 | 2 | 0.767 |
| Aplastic R. Olf. Bulb | 69 | **30** | 2 | **<0.001** | 49 | 2 | 28 | 0.281 |
| Hypoplastic R. Olf. Bulb | 65 | 4 | 3 | 0.515 | 45 | 0 | 4 | 0.660 |
| Combination R. Olf. Bulb | 69 | **34** | 5 | **<0.001** | 49 | 2 | 32 | 0.301 |
| Dysplastic R. Olf. Bulb | 65 | 0 | 0 | . | 45 | 0 | 0 | . |
| Hypoplastic Brainstem | 66 | 0 | 0 | . | 46 | 0 | 0 | . |
| Dysplastic Brainstem | 67 | **18** | 2 | **0.016** | 47 | 2 | 16 | 0.069 |
| Hypoplastic Midbrain | 67 | 0 | 0 | . | 47 | 0 | 0 | . |
| Dysplastic Midbrain | 67 | **17** | 1 | **0.006** | 47 | 2 | 15 | 0.057 |
| BDS Dichotomized | 69 | **45** | 10 | **<0.001** | 49 | 2 | 43 | 0.635 |
| BDS Hippocampus or Cerebellum | 69 | **45** | 10 | **<0.001** | 49 | 2 | 43 | 0.675 |

* *P value is calculated from two sample Student T-test*

**Supplemental Table 6B: Brain Dysplasia Score within Ohia Mouse Mutant Cohort: Comparison between Cardiac Lesion Subgroups**

|  | Conotruncal vs. Non Conotruncal | | | | Cyanotic vs. Acyanotic | | | | Arch Obstruction vs. No Arch Obstruction | | | |
| --- | --- | --- | --- | --- | --- | --- | --- | --- | --- | --- | --- | --- |
| Structure | N | Cono. | Non Cono. | P value * | N | Cyanotic | Acyanotic | P value * | N | Obstr. | No Obstr. | P value * |
| Aplastic Hippocampus | 49 | 3 | 5 | 0.723 | 49 | 3 | 5 | 0.839 | 49 | 6 | 2 | 0.144 |
| Hypoplastic Hippocampus | 48 | 2 | 8 | 0.425 | 48 | 4 | 6 | 0.977 | 48 | 4 | 6 | 0.488 |
| Combination Hippocampus | 49 | 14 | 29 | 0.900 | 49 | 18 | 25 | 0.819 | 49 | 22 | 21 | 0.313 |
| Dysplastic Hippocampus | 48 | **10** | 23 | 0.948 | 48 | 14 | 19 | 0.560 | 48 | 15 | 18 | 0.361 |
| Hypoplastic Cerebrum | 48 | **1** | 12 | **0.037** | 48 | 2 | 11 | **0.037** | 48 | 7 | 6 | 0.752 |
| Dysplastic Cerebrum | 48 | **10** | 25 | 0.668 | 48 | 13 | 22 | 0.828 | 48 | 18 | 17 | 0.365 |
| Aplastic Cerebellum | 49 | 0 | 1 | 0.498 | 49 | 1 | 0 | 0.232 | 49 | 1 | 0 | 0.332 |
| Hypoplastic Cerebellum | 47 | 2 | 4 | 0.909 | 47 | 3 | 3 | 0.618 | 47 | 2 | 4 | 0.424 |
| Combination Cerebellum | 49 | 13 | 29 | 0.692 | 49 | 17 | **25** | 0.240 | 49 | 23 | **19** | 0.957 |
| Dysplastic Cerebellum | 47 | **12** | 28 | 0.684 | 47 | 15 | 25 | 0.339 | 47 | 22 | 18 | **0.048** |
| Aplastic L. Olf. Bulb | 49 | **12** | 18 | 0.143 | 49 | 15 | 15 | 0.104 | 49 | 16 | 14 | 0.692 |
| Hypoplastic L. Olf. Bulb | 47 | 0 | 7 | 0.055 | 47 | 0 | 7 | **0.018** | 47 | 4 | 3 | 0.647 |
| Combination L. Olf. Bulb | 49 | **12** | 25 | 0.379 | 49 | 15 | 22 | 0.375 | 49 | 20 | 17 | 0.551 |
| Dysplastic L. Olf. Bulb | 47 | 0 | 2 | 0.341 | 47 | 0 | 2 | 0.243 | 47 | 1 | 1 | 0.976 |
| Aplastic R. Olf. Bulb | 49 | **12** | 18 | 0.202 | 49 | 15 | 15 | 0.164 | 49 | 16 | 14 | 0.493 |
| Hypoplastic R. Olf. Bulb | 45 | 0 | 4 | 0.152 | 45 | 0 | 4 | 0.076 | 45 | 2 | 2 | 0.964 |
| Combination R. Olf. Bulb | 49 | **12** | 22 | 0.344 | 49 | 15 | 19 | 0.329 | 49 | 18 | 16 | 0.467 |
| Dysplastic R. Olf. Bulb | 45 | 0 | 0 | . | 45 | 0 | 0 | . | 45 | 0 | 0 | . |
| Hypoplastic Brainstem | 46 | 0 | 0 | . | 46 | 0 | 0 | . | 46 | 0 | 0 | . |
| Dysplastic Brainstem | 47 | **6** | 12 | 0.814 | 47 | 10 | 8 | 0.100 | 47 | 11 | 7 | 0.196 |
| Hypoplastic Midbrain | 47 | 0 | 0 | . | 47 | 0 | 0 | . | 47 | 0 | 0 | . |
| Dysplastic Midbrain | 47 | **7** | 10 | 0.282 | 47 | 10 | 7 | 0.055 | 47 | 9 | 8 | 0.687 |
| BDS Dichotomized | 49 | **14** | 31 | 0.942 | 49 | 18 | 27 | 0.970 | 49 | 24 | 21 | 0.612 |
| BDS Hipp. or Cerebellum | 49 | **14** | 31 | 0.693 | 49 | 18 | 27 | 0.704 | 49 | 24 | 21 | 0.287 |

* *Two Sample Student T-test used to examine incidence differences between groupings.*

**Supplemental Table 8A: Non-normalized Regional Cerebral Volumes of Ohia CHD cohorts: Comparison of Cardiac Lesion Subgroups**

|  | CHD vs. No-CHD | | | | | Single vs. Biventricular Morphology | | | | |
| --- | --- | --- | --- | --- | --- | --- | --- | --- | --- | --- |
|  | CHD | | No CHD | | P value * | Single | | Biventricular | | P value * |
| Structure | N | Mean (x10^8^ μm^3^) | N | Mean (x10^8^ μm^3^) |  | N | Mean (x10^8^ μm^3^) | N | Mean (x10^8^ μm^3^) |  |
| R. Hippocampus | 7 | 0.72 | 11 | 0.96 | 0.340 | 1 | 0.07 | 10 | 0.79 | . |
| R. Olf. Bulb | 6 | 1.93 | 9 | 2.36 | 0.500 | 0 | . | 9 | 1.93 | . |
| R. Subcortical | 7 | 3.59 | 16 | 4.97 | 0.237 | 2 | 2.45 | 14 | **3.76** | **0.006** |
| R. Cortex | 7 | 6.99 | 14 | 10.58 | 0.188 | 1 | 2.43 | 13 | 7.34 | . |
| Intraventricular Vol. | 2 | 2.10 | 8 | 1.70 | 0.535 | 1 | 2.29 | 7 | 2.07 | . |
| Midbrain | 7 | 5.60 | 16 | 6.53 | 0.597 | 2 | 4.67 | 14 | 5.73 | 0.192 |
| L. Hippocampus | 7 | 0.76 | 12 | 1.05 | 0.252 | 1 | 0.03 | 11 | 0.82 | . |
| L. Olf. Bulb | 6 | 1.79 | 10 | 2.52 | 0.314 | 0 | . | 10 | 1.79 | . |
| L. Subcortical | 7 | 3.65 | 16 | 5.00 | 0.242 | 2 | 2.79 | 14 | **3.78** | **0.036** |
| L. Cortex | 7 | 7.08 | 15 | 10.64 | 0.197 | 1 | 2.72 | 14 | 7.39 | . |
| Cerebellum | 7 | 1.61 | 15 | 1.61 | 1.000 | 1 | 1.00 | 14 | 1.65 | . |
| Pons | 7 | 2.08 | 16 | 2.50 | 0.572 | 2 | 1.20 | 14 | **2.21** | **0.024** |
| Medulla | 7 | 5.93 | 15 | 7.47 | 0.319 | 1 | 5.76 | 14 | 5.95 | . |
| Hypothalamus | 3 | 0.14 | 3 | 0.09 | 0.456 | 0 | . | 3 | 0.14 | . |
| Choroid Plexus | 6 | 0.26 | 16 | 0.21 | 0.530 | 2 | 0.32 | 14 | 0.25 | 0.743 |
| Supratentorial | 7 | 30.18 | 16 | 44.59 | 0.207 | 2 | 14.00 | 14 | 32.49 | 0.057 |
| Infratentorial | 7 | 9.18 | 16 | 11.62 | 0.364 | 2 | 4.58 | 14 | 9.83 | 0.374 |
| Total Volume | 7 | 40.65 | 16 | 57.36 | 0.230 | 2 | 21.18 | 14 | 43.43 | 0.247 |

** Welch–Satterthwaite t-test was used to test volume differences among individual structures between groups.*

**Supplemental Table 8B: Non-normalized Regional Cerebral Volumes of Ohia CHD cohorts: Comparison of Cardiac Lesion Subgroups**

|  | Conotruncal vs. Non-Conotruncal | | | | | Cyanotic vs. Acyanotic Lesion | | | | | Arch Obstruction vs. No Arch Obstruction | | | | |
| --- | --- | --- | --- | --- | --- | --- | --- | --- | --- | --- | --- | --- | --- | --- | --- |
|  | Conotruncal | | Non Conotruncal | | P value * | Cyanotic | | Acyanotic | | P value * | Arch Obstruction | | No Obstruction | | P value * |
| Structure | N | Mean (x10^8^ μm^3^) | N | Mean (x10^8^ μm^3^) |  | N | Mean (x10^8^ μm^3^) | N | Mean (x10^8^ μm^3^) |  | N | Mean (x10^8^ μm^3^) | N | Mean (x10^8^ μm^3^) |  |
| R. Hippocampus | 3 | 0.49 | 9 | 0.79 | 0.117 | 5 | 0.53 | 6 | 0.89 | 0.114 | 4 | 0.80 | 7 | 0.68 | 0.513 |
| R. Olf. Bulb | 2 | 1.47 | 8 | 1.94 | 0.159 | 3 | 1.51 | 6 | 2.14 | 0.121 | 4 | 1.66 | 5 | 2.14 | 0.325 |
| R. Subcortical | 4 | 2.87 | 13 | **3.77** | **0.043** | 7 | 2.96 | 9 | 4.08 | 0.057 | 7 | 3.54 | 9 | 3.63 | 0.889 |
| R. Cortex | 3 | 5.88 | 12 | 7.26 | 0.412 | 5 | 5.57 | 9 | 7.78 | 0.249 | 6 | 6.03 | 8 | 7.71 | 0.419 |
| Intraventricular Vol. | 2 | 2.10 | 6 | 2.10 | 0.999 | 3 | 2.16 | 5 | 2.06 | 0.944 | 3 | 1.69 | 5 | 2.34 | 0.643 |
| Midbrain | 4 | 5.22 | 13 | 5.61 | 0.675 | 7 | 5.02 | 9 | 6.05 | 0.398 | 7 | 4.62 | 9 | 6.36 | 0.159 |
| L. Hippocampus | 4 | 0.59 | 9 | 0.82 | 0.228 | 6 | 0.55 | 6 | **0.97** | **0.044** | 4 | 0.81 | 8 | 0.73 | 0.679 |
| L. Olf. Bulb | 3 | 1.60 | 8 | 1.79 | 0.556 | 4 | 1.56 | 6 | 1.95 | 0.340 | 4 | 1.39 | 6 | 2.06 | 0.097 |
| L. Subcortical | 4 | 3.11 | 13 | 3.78 | 0.141 | 7 | 3.22 | 9 | 3.99 | 0.211 | 7 | 3.40 | 9 | 3.85 | 0.496 |
| L. Cortex | 4 | 6.43 | 12 | 7.30 | 0.591 | 6 | 6.22 | 9 | 7.66 | 0.434 | 6 | 5.98 | 9 | 7.81 | 0.334 |
| Cerebellum | 4 | 1.56 | 12 | 1.61 | 0.811 | 6 | 1.54 | 9 | 1.65 | 0.657 | 6 | 1.45 | 9 | 1.71 | 0.313 |
| Pons | 4 | 1.99 | 13 | 2.07 | 0.788 | 7 | 1.89 | 9 | 2.23 | 0.429 | 7 | 1.65 | 9 | 2.42 | 0.073 |
| Medulla | 4 | 5.69 | 12 | 6.13 | 0.493 | 6 | 6.33 | 9 | 5.67 | 0.473 | 6 | 5.87 | 9 | 5.98 | 0.913 |
| Hypothalamus | 1 | 0.11 | 3 | 0.12 | . | 2 | 0.09 | 1 | 0.23 | . | 0 | . | 3 | 0.14 | . |
| Choroid Plexus | 4 | 0.24 | 13 | 0.26 | 0.878 | 7 | 0.25 | 9 | 0.26 | 0.936 | 7 | 0.31 | 9 | 0.22 | 0.579 |
| Supratentorial | 4 | 26.22 | 13 | 31.25 | 0.344 | 7 | 24.06 | 9 | 34.93 | 0.138 | 7 | 25.55 | 9 | 33.77 | 0.285 |
| Infratentorial | 4 | 9.27 | 13 | 9.24 | 0.98 | 7 | 8.66 | 9 | 9.58 | 0.610 | 7 | 7.92 | 9 | 10.16 | 0.213 |
| Total Volume | 4 | 35.58 | 13 | 42.06 | 0.314 | 7 | 33.52 | 9 | 46.19 | 0.163 | 7 | 34.31 | 9 | 45.57 | 0.238 |

** Welch–Satterthwaite t-test was used to test volume differences among individual structures between groups.*

**Supplemental Table 9A: Normalized Regional Cerebral Volumes of Ohia CHD cohorts: Comparison of Cardiac Lesion Subgroups**

|  | CHD vs. No-CHD | | | Single vs.Biventricular Morphology | | |
| --- | --- | --- | --- | --- | --- | --- |
| Structure | CHD Percent TBV | No CHD Percent TBV | P value * | Single Percent TBV | Bivent. Percent TBV | P value * |
| R. Hippocampus | 0.015 | 0.017 | 0.376 | 0.002 | 0.016 | . |
| R. Olf. Bulb | 0.037 | 0.039 | 0.709 | . | 0.037 | . |
| R. Subcortical | 0.099 | 0.086 | 0.198 | 0.150 | 0.092 | 0.5485 |
| R. Cortex | **0.150** | 0.183 | **0.026** | 0.076 | 0.156 | . |
| Intraventricular Vol. | 0.049 | 0.026 | 0.155 | 0.072 | 0.046 | . |
| Midbrain | **0.155** | 0.111 | **0.044** | 0.289 | 0.136 | 0.4591 |
| L. Hippocampus | 0.016 | 0.019 | 0.262 | 0.001 | 0.018 | . |
| L. Olf. Bulb | 0.037 | 0.040 | 0.521 | . | 0.037 | . |
| L. Subcortical | 0.102 | 0.087 | 0.206 | 0.171 | 0.092 | 0.4917 |
| L. Cortex | 0.159 | 0.184 | 0.106 | 0.085 | 0.164 | . |
| Cerebellum | **0.041** | 0.030 | **0.044** | 0.031 | 0.042 | . |
| Pons | 0.055 | 0.046 | 0.292 | 0.070 | 0.053 | 0.6264 |
| Medulla | 0.155 | 0.141 | 0.516 | 0.180 | 0.153 | . |
| Hypothalamus | 0.002 | 0.002 | 0.611 | . | 0.002 | . |
| Choroid Plexus | 0.007 | 0.004 | 0.057 | **0.015** | 0.006 | **0.0003** |
| Supratentorial | 0.735 | 0.764 | 0.324 | 0.743 | 0.733 | 0.963 |
| Infratentorial | 0.239 | 0.218 | 0.506 | 0.176 | 0.248 | 0.5293 |

** Welch–Satterthwaite t-test was used to test percent volume differences among individual structures between groups.*

|  | Conotruncal vs. Non-Conotruncal | | | Cyanotic vs. Acyanotic Lesion | | | Arch Obstruction vs. No Arch Obstruction | | |
| --- | --- | --- | --- | --- | --- | --- | --- | --- | --- |
| Structure | Cono. Percent TBV | Non Cono. Percent TBV | P value * | Cyanotic Percent TBV | Acyanotic Percent TBV | P value * | Obstr. Percent TBV | No Obstr. Percent TBV | P value * |
| R. Hippocampus | 0.013 | 0.016 | 0.412 | 0.013 | 0.017 | 0.293 | 0.019 | 0.013 | 0.097 |
| R. Olf. Bulb | 0.037 | 0.036 | 0.820 | 0.035 | 0.038 | 0.329 | 0.037 | 0.037 | 0.937 |
| R. Subcortical | 0.083 | 0.103 | 0.217 | 0.103 | 0.096 | 0.783 | 0.121 | 0.082 | 0.086 |
| R. Cortex | 0.155 | 0.151 | 0.890 | 0.139 | 0.156 | 0.517 | 0.145 | 0.154 | 0.760 |
| Intraventricular Vol. | 0.064 | 0.044 | 0.809 | 0.067 | 0.039 | 0.532 | 0.036 | 0.057 | 0.470 |
| Midbrain | 0.146 | 0.154 | 0.756 | 0.180 | 0.135 | 0.328 | 0.171 | 0.142 | 0.529 |
| L. Hippocampus | 0.017 | 0.016 | 0.876 | 0.014 | 0.018 | 0.377 | 0.019 | 0.015 | 0.242 |
| L. Olf. Bulb | 0.044 | 0.034 | 0.079 | 0.040 | 0.035 | 0.344 | 0.032 | 0.040 | 0.075 |
| L. Subcortical | 0.089 | 0.104 | 0.413 | 0.113 | 0.093 | 0.440 | 0.120 | 0.087 | 0.211 |
| L. Cortex | 0.183 | 0.153 | 0.477 | 0.166 | 0.155 | 0.739 | 0.144 | 0.169 | 0.402 |
| Cerebellum | 0.044 | 0.040 | 0.347 | 0.041 | 0.041 | 0.978 | 0.044 | 0.039 | 0.624 |
| Pons | 0.056 | 0.054 | 0.643 | 0.060 | 0.051 | 0.241 | 0.056 | 0.054 | 0.821 |
| Medulla | 0.162 | 0.155 | 0.735 | 0.170 | 0.145 | 0.289 | 0.166 | 0.147 | 0.498 |
| Hypothalamus | 0.003 | 0.002 | . | 0.002 | 0.003 | . | . | 0.002 | . |
| Choroid Plexus | 0.007 | 0.007 | 0.919 | 0.009 | 0.006 | 0.510 | 0.009 | 0.006 | 0.374 |
| Supratentorial | 0.734 | 0.734 | 0.997 | 0.734 | 0.735 | 0.969 | 0.746 | 0.726 | 0.665 |
| Infratentorial | 0.263 | 0.234 | 0.278 | 0.241 | 0.237 | 0.900 | 0.236 | 0.241 | 0.907 |

**Supplemental Table 9B: Normalized Regional Cerebral Volumes of Ohia CHD cohorts: Comparison of Cardiac Lesion Subgroups**

** Welch–Satterthwaite t-test was used to test percent volume differences among individual structures between groups.*

**Supplemental Table 10: OVERVIEW OF OHIA MUTANT MODEL GENOTYPE GROUPS STUDIES: CHD lesions and Holoprosencephaly Incidence**

|  | A^†^ | B^‡^ | C^§^ | D^\|\|^ | E^¶^ | F^#^ |
| --- | --- | --- | --- | --- | --- | --- |
| N Total | 24 | 8 | 7 | 4 | 0 | 20 |
| CHD | 21 | 7 | 6 | 3 | 0 | 10 |
| Holoprosencephaly | 19 | 6 | 4 | 4 | 0 | 8 |
| Conotruncal | 7 | 3 | 0 | 1 | 0 | 5 |
| Single Ventricle | 1 | 1 | 0 | 0 | 0 | 0 |
| Cyanosis | 8 | 4 | 1 | 1 | 0 | 6 |
| ArchObst | 16 | 1 | 3 | 2 | 0 | 2 |

†Group A: N 24, Genotype: Pcdha9(m/m) Sap130(m/m)

‡ Group B: N 8, Genotype: Pcdha9(m/+) Sap130(m/m)

§ Group C: N 7, Genotype: Pcdha9(+/+) Sap130(m/m)

|| Group D: N 4, Genotype: Pcdha9(m/m) Sap130(m/+)

¶ Group E: N 0, Genotype: Pcdha9(m/m) Sap130(+/+)

\# Group F: N 27, Genotype: Pcdha9(m/+) or (+/+) Sap130(m/+) or (+/+)

**Supplemental Table 11A: Genotype/Brain Dysplasia Score : Sap130(m/m), Pcdha9 (*/*) vs WT; Sap130(m/m), Pcdha9 (*/*) vs Sap130(*/+), Pcdha9 (*/+)**

|  | ABC vs. WT | | | ABC vs. F | | |
| --- | --- | --- | --- | --- | --- | --- |
|  | Sap130(m/m), Pcdha9 (*/*) vs WT | | | Sap130(m/m), Pcdha9 (*/*) vs Sap130(*/+), Pcdha9 (*/+) | | |
| Structure | ABC Incidence | WT Incidence | P value * | ABC Incidence | F Incidence | P value * |
| **Aplastic Hippocampus** | 05/39 (12.82%) | 0/10 (0.00%) | 0.232 | 05/39 (12.82%) | 01/20 (5.00%) | 0.347 |
| **Hypoplastic Hippocampus** | 06/38 (15.79%) | 0/10 (0.00%) | 0.179 | 06/38 (15.79%) | 05/20 (25.00%) | 0.395 |
| **Combination Hippocampus** | 11/39 (28.21%) | 0/10 (0.00%) | 0.057 | 11/39 (28.21%) | 06/20 (30.00%) | 0.885 |
| **Dysplastic Hippocampus** | **27/38 (71.05%)** | 0/10 (0.00%) | **<.001** | 27/38 (71.05%) | 12/20 (60.00%) | 0.394 |
| **Hypoplastic Cerebrum** | **12/38 (31.58%)** | 0/10 (0.00%) | **0.040** | **12/38 (31.58%)** | 01/20 (5.00%) | **0.021** |
| **Dysplastic Cerebrum** | **29/38 (76.32%)** | 0/10 (0.00%) | **<.001** | **29/38 (76.32%)** | 08/20 (40.00%) | **0.016** |
| **Aplastic Cerebellum** | 01/39 (2.56%) | 0/10 (0.00%) | 0.609 | 01/39 (2.56%) | 01/20 (5.00%) | 0.625 |
| **Hypoplastic Cerebellum** | 04/37 (10.81%) | 0/10 (0.00%) | 0.277 | 04/37 (10.81%) | 05/20 (25.00%) | 0.161 |
| **Combination Cerebellum** | 05/39 (12.82%) | 0/10 (0.00%) | 0.232 | 05/39 (12.82%) | 06/20 (30.00%) | 0.109 |
| **Dysplastic Cerebellum** | **34/37 (91.89%)** | 0/10 (0.00%) | **<.001** | **34/37 (91.89%)** | 09/20 (45.00%) | **<.001** |
| **Aplastic L. Olf. Bulb** | **23/39 (58.97%)** | 0/10 (0.00%) | **0.001** | **23/39 (58.97%)** | 06/20 (30.00%) | **0.035** |
| **Hypoplastic L. Olf. Bulb** | 06/37 (16.22%) | 0/10 (0.00%) | 0.173 | 06/37 (16.22%) | 02/20 (10.00%) | 0.519 |
| **Combination L. Olf. Bulb** | **29/39 (74.36%)** | 0/10 (0.00%) | **<.001** | **29/39 (74.36%)** | 08/20 (40.00%) | **0.010** |
| **Dysplastic L. Olf. Bulb** | 01/37 (2.70%) | 0/10 (0.00%) | 0.599 | 01/37 (2.70%) | 01/20 (5.00%) | 0.653 |
| **Aplastic R. Olf. Bulb** | **24/39 (61.54%)** | 0/10 (0.00%) | **0.001** | **24/39 (61.54%)** | 06/20 (30.00%) | **0.022** |
| **Hypoplastic R. Olf. Bulb** | 04/36 (11.11%) | 0/10 (0.00%) | 0.270 | 04/36 (11.11%) | 02/19 (10.53%) | 0.947 |
| **Combination R. Olf. Bulb** | **28/39 (71.79%)** | 0/10 (0.00%) | **<.001** | **28/39 (71.79%)** | 08/20 (40.00%) | **0.018** |
| **Dysplastic R. Olf. Bulb** | 00/36 (0.00%) | 0/10 (0.00%) | . | 00/36 (0.00%) | 00/19 (0.00%) | . |
| **Hypoplastic Brainstem** | 00/37 (0.00%) | 0/10 (0.00%) | . | 00/37 (0.00%) | 00/19 (0.00%) | . |
| **Dysplastic Brainstem** | **12/37 (32.43%)** | 0/10 (0.00%) | **0.037** | 12/37 (32.43%) | 06/20 (30.00%) | 0.850 |
| **Hypoplastic Midbrain** | 00/37 (0.00%) | 0/10 (0.00%) | . | 00/37 (0.00%) | 00/20 (0.00%) | . |
| **Dysplastic Midbrain** | 10/37 (27.03%) | 0/10 (0.00%) | 0.064 | 10/37 (27.03%) | 06/20 (30.00%) | 0.812 |
| **BDS Dichotomized** | **35/39 (89.74%)** | 0/10 (0.00%) | **<.001** | **35/39 (89.74%)** | 13/20 (65.00%) | **0.021** |

** Chi-square analysis was used to compare individual structures and BDS Dichotomized.*

**Supplemental Table 11B: Genotype/Brain Dysplasia Score: Sap130(m/m), Pcdha9 (m/*) vs Sap130(*/+), Pcdha9 (*/+); Sap130(m/m), Pcdha9 (m/m) vs Sap130(m/m), Pcdha9 (+/+)**

|  | AB vs. F | | | A vs. C | | |
| --- | --- | --- | --- | --- | --- | --- |
|  | Sap130(m/m), Pcdha9 (m/*) vs Sap130(*/+), Pcdha9 (*/+) | | | Sap130(m/m), Pcdha9 (m/m) vs Sap130(m/m), Pcdha9 (+/+) | | |
| Structure | AB Incidence | F Incidence | P value * | A Incidence | C Incidence | P value * |
| **Aplastic Hippocampus** | 05/32 (15.63%) | 01/20 (5.00%) | 0.243 | 05/24 (20.83%) | 00/7 (0.00%) | 0.187 |
| **Hypoplastic Hippocampus** | 06/31 (19.35%) | 05/20 (25.00%) | 0.632 | 04/23 (17.39%) | 00/7 (0.00%) | 0.236 |
| **Combination Hippocampus** | 11/32 (34.38%) | 06/20 (30.00%) | 0.744 | 09/24 (37.50%) | 00/7 (0.00%) | 0.054 |
| **Dysplastic Hippocampus** | 21/31 (67.74%) | 12/20 (60.00%) | 0.572 | 15/23 (65.22%) | 06/7 (85.71%) | 0.300 |
| **Hypoplastic Cerebrum** | **09/31 (29.03%)** | 01/20 (5.00%) | **0.035** | 07/23 (30.43%) | 03/7 (42.86%) | 0.542 |
| **Dysplastic Cerebrum** | **24/31 (77.42%)** | 08/20 (40.00%) | **0.019** | 18/23 (78.26%) | 05/7 (71.43%) | 0.708 |
| **Aplastic Cerebellum** | 01/32 (3.13%) | 01/20 (5.00%) | 0.732 | 01/24 (4.17%) | 00/7 (0.00%) | 0.583 |
| **Hypoplastic Cerebellum** | 04/30 (13.33%) | 05/20 (25.00%) | 0.293 | 02/22 (9.09%) | 00/7 (0.00%) | 0.408 |
| **Combination Cerebellum** | 05/32 (15.63%) | 06/20 (30.00%) | 0.217 | 03/24 (12.50%) | 00/7 (0.00%) | 0.325 |
| **Dysplastic Cerebellum** | **27/30 (90.00%)** | 09/20 (45.00%) | **0.001** | 20/22 (90.91%) | 07/7 (100.00%) | 0.408 |
| **Aplastic L. Olf. Bulb** | **19/32 (59.38%)** | 06/20 (30.00%) | **0.039** | 14/24 (58.33%) | 04/7 (57.14%) | 0.955 |
| **Hypoplastic L. Olf. Bulb** | 04/30 (13.33%) | 02/20 (10.00%) | 0.722 | 03/22 (13.64%) | 02/7 (28.57%) | 0.362 |
| **Combination L. Olf. Bulb** | **23/32 (71.88%)** | 08/20 (40.00%) | **0.023** | 17/24 (70.83%) | 06/7 (85.71%) | 0.429 |
| **Dysplastic L. Olf. Bulb** | 01/30 (3.33%) | 01/20 (5.00%) | 0.768 | 01/22 (4.55%) | 00/7 (0.00%) | 0.566 |
| **Aplastic R. Olf. Bulb** | **20/32 (62.50%)** | 06/20 (30.00%) | **0.023** | 15/24 (62.50%) | 04/7 (57.14%) | 0.798 |
| **Hypoplastic R. Olf. Bulb** | 03/29 (10.34%) | 02/19 (10.53%) | 0.984 | 02/21 (9.52%) | 01/7 (14.29%) | 0.724 |
| **Combination R. Olf. Bulb** | **23/32 (71.88%)** | 08/20 (40.00%) | **0.023** | 17/24 (70.83%) | 05/7 (71.43%) | 0.976 |
| **Dysplastic R. Olf. Bulb** | 00/29 (0.00%) | 00/19 (0.00%) | . | 00/21 (0.00%) | 00/7 (0.00%) | . |
| **Hypoplastic Brainstem** | 00/30 (0.00%) | 00/19 (0.00%) | . | 00/22 (0.00%) | 00/7 (0.00%) | . |
| **Dysplastic Brainstem** | 12/30 (40.00%) | 06/20 (30.00%) | 0.471 | **10/22 (45.45%)** | 00/7 (0.00%) | **0.028** |
| **Hypoplastic Midbrain** | 00/30 (0.00%) | 00/20 (0.00%) | . | 00/22 (0.00%) | 00/7 (0.00%) | . |
| **Dysplastic Midbrain** | 10/30 (33.33%) | 06/20 (30.00%) | 0.805 | **09/22 (40.91%)** | 00/7 (0.00%) | **0.042** |
| **BDS Dichotomized** | 28/32 (87.50%) | 13/20 (65.00%) | 0.053 | 21/24 (87.50%) | 07/7 (100.00%) | 0.325 |

** Chi-square analysis was used to compare individual structures and BDS Dichotomized*

**Supplemental Table 11C: Genotype/Brain Dysplasia Score: Sap130(m/m), Pcdha9 (m/+) vs Sap130(m/+), Pcdha9 (m/m)**

|  | B vs. D | | |
| --- | --- | --- | --- |
|  | Sap130(m/m), Pcdha9 (m/+) vs Sap130(m/+), Pcdha9 (m/m) | | |
| Structure | B Incidence | D Incidence | P value * |
| Aplastic Hippocampus | 00/8 (0.00%) | **02/4 (50.00%)** | **0.029** |
| Hypoplastic Hippocampus | 02/8 (25.00%) | 02/4 (50.00%) | 0.387 |
| Combination Hippocampus | 02/8 (25.00%) | **04/4 (100.00%)** | **0.014** |
| Dysplastic Hippocampus | 06/8 (75.00%) | 02/4 (50.00%) | 0.387 |
| Hypoplastic Cerebrum | 02/8 (25.00%) | 00/4 (0.00%) | 0.273 |
| Dysplastic Cerebrum | 06/8 (75.00%) | 04/4 (100.00%) | 0.273 |
| Aplastic Cerebellum | 00/8 (0.00%) | 00/4 (0.00%) | . |
| Hypoplastic Cerebellum | 02/8 (25.00%) | 00/4 (0.00%) | 0.273 |
| Combination Cerebellum | 02/8 (25.00%) | 00/4 (0.00%) | 0.273 |
| Dysplastic Cerebellum | 07/8 (87.50%) | 04/4 (100.00%) | 0.460 |
| Aplastic L. Olf. Bulb | 05/8 (62.50%) | 03/4 (75.00%) | 0.665 |
| Hypoplastic L. Olf. Bulb | 01/8 (12.50%) | 01/4 (25.00%) | 0.584 |
| Combination L. Olf. Bulb | 06/8 (75.00%) | 04/4 (100.00%) | 0.273 |
| Dysplastic L. Olf. Bulb | 00/8 (0.00%) | 00/4 (0.00%) | . |
| Aplastic R. Olf. Bulb | 05/8 (62.50%) | 03/4 (75.00%) | 0.665 |
| Hypoplastic R. Olf. Bulb | 01/8 (12.50%) | 01/4 (25.00%) | 0.584 |
| Combination R. Olf. Bulb | 06/8 (75.00%) | 04/4 (100.00%) | 0.273 |
| Dysplastic R. Olf. Bulb | 00/8 (0.00%) | 00/4 (0.00%) | . |
| Hypoplastic Brainstem | 00/8 (0.00%) | 00/4 (0.00%) | . |
| Dysplastic Brainstem | 02/8 (25.00%) | 02/4 (50.00%) | 0.387 |
| Hypoplastic Midbrain | 00/8 (0.00%) | 00/4 (0.00%) | . |
| Dysplastic Midbrain | 01/8 (12.50%) | 02/4 (50.00%) | 0.157 |
| BDS Dichotomized | 07/8 (87.50%) | 04/4 (100.00%) | 0.460 |

** Welch–Satterthwaite t-test was used to test volume differences among individual structures between groups.*

**Supplemental Table 12A: Genotype/Regional Brain Volumes: Sap130(m/m), Pcdha9 (*/*) vs WT; Sap130(m/m), Pcdha9 (*/*) vs Sap130(*/+), Pcdha9 (*/+)**

|  | ABC vs. WT | | | | | ABC vs. F | | | | |
| --- | --- | --- | --- | --- | --- | --- | --- | --- | --- | --- |
|  | Sap130(m/m), Pcdha9 (*/*) vs WT | | | | | Sap130(m/m), Pcdha9 (*/*) vs Sap130(*/+), Pcdha9 (*/+) | | | | |
|  | ABC | | WT | | P value * | ABC | | F | | P value * |
| Structure | N | Mean (x10^8^ μm^3^) | N | Mean (x10^8^ μm^3^) |  | N | Mean (x10^8^ μm^3^) | N | Mean (x10^8^ μm^3^) |  |
| R. Hippocampus | 9 | 0.61 | 10 | 0.98 | 0.077 | 9 | 0.61 | 9 | **1.03** | **0.041** |
| R. Olf. Bulb | 7 | 1.63 | 10 | 2.19 | 0.171 | 7 | 1.63 | 8 | 2.50 | 0.096 |
| R. Subcortical | 14 | 3.31 | 10 | **5.80** | **0.006** | 14 | 3.31 | 9 | **5.06** | **0.050** |
| R. Cortex | 12 | 6.00 | 10 | **9.39** | **0.047** | 12 | 6.00 | 9 | **11.26** | **0.016** |
| Intraventricular Vol. | 7 | 1.81 | 9 | 0.53 | 0.083 | 7 | 1.81 | 3 | 2.50 | 0.530 |
| Midbrain | 14 | 4.70 | 10 | 5.35 | 0.397 | 14 | 4.70 | 9 | 7.57 | 0.078 |
| L. Hippocampus | 10 | 0.63 | 10 | 1.04 | 0.051 | 10 | 0.63 | 9 | **1.09** | **0.031** |
| L. Olf. Bulb | 8 | 1.52 | 10 | 2.21 | 0.085 | 8 | 1.52 | 8 | **2.60** | **0.048** |
| L. Subcortical | 14 | 3.39 | 10 | **5.88** | **0.007** | 14 | 3.39 | 9 | 5.04 | 0.062 |
| L. Cortex | 13 | 6.15 | 10 | 9.13 | 0.061 | 13 | 6.15 | 9 | **11.13** | **0.022** |
| Cerebellum | 13 | 1.53 | 9 | **2.45** | **0.003** | 13 | 1.53 | 9 | 1.74 | 0.516 |
| Pons | 14 | 1.84 | 10 | 2.66 | 0.106 | 14 | 1.84 | 9 | 2.74 | 0.144 |
| Medulla | 13 | 5.82 | 10 | 6.80 | 0.414 | 13 | 5.82 | 9 | 7.13 | 0.291 |
| Hypothalamus | 1 | 0.07 | 10 | 0.15 | . | 1 | 0.07 | 5 | 0.12 | . |
| Choroid Plexus | 14 | 0.27 | 10 | 0.23 | 0.647 | 14 | 0.27 | 8 | 0.18 | 0.272 |
| Supratentorial | 14 | 25.95 | 10 | **68.46** | **0.013** | 14 | 25.95 | 9 | **47.72** | **0.018** |
| Infratentorial | 14 | 8.66 | 10 | 11.81 | 0.128 | 14 | 8.66 | 9 | 11.68 | 0.149 |
| Total Volume | 14 | 35.49 | 10 | **54.50** | **0.028** | 14 | 35.49 | 9 | **61.23** | **0.022** |

** Welch–Satterthwaite t-test was used to test volume differences among individual structures between groups.*

**Supplemental Table 12B: Genotype/Regional Brain Volumes: Sap130(m/m), Pcdha9 (m/*) vs Sap130(*/+), Pcdha9 (*/+);Sap130(m/m), Pcdha9 (m/m) vs Sap130(m/m), Pcdha9 (+/+)**

|  | AB vs. F | | | | | A vs. C | | | | |
| --- | --- | --- | --- | --- | --- | --- | --- | --- | --- | --- |
|  | Sap130(m/m), Pcdha9 (m/*) vs Sap130(*/+), Pcdha9 (*/+) | | | | | Sap130(m/m), Pcdha9 (m/m) vs Sap130(m/m), Pcdha9 (+/+) | | | | |
|  | AB | | F | | P value * | A | | C | | P value * |
| Structure | N | Mean (x10^8^ μm^3^) | N | Mean (x10^8^ μm^3^) |  | N | Mean (x10^8^ μm^3^) | N | Mean (x10^8^ μm^3^) |  |
| R. Hippocampus | 7 | 0.52 | 9 | **1.03** | **0.021** | 3 | 0.56 | 2 | 0.90 | 0.281 |
| R. Olf. Bulb | 5 | 1.54 | 8 | 2.50 | 0.121 | 3 | 1.16 | 2 | 1.87 | 0.195 |
| R. Subcortical | 10 | 2.94 | 9 | **5.06** | **0.023** | 5 | 2.27 | 4 | **4.24** | **0.022** |
| R. Cortex | 8 | 5.79 | 9 | **11.26** | **0.029** | 4 | 4.72 | 4 | 6.41 | 0.423 |
| Intraventricular Vol. | 4 | 1.64 | 3 | 2.50 | 0.532 | 2 | 0.05 | 3 | 2.04 | 0.128 |
| Midbrain | 10 | 4.48 | 9 | 7.57 | 0.064 | 5 | 3.88 | 4 | 5.26 | 0.193 |
| L. Hippocampus | 8 | 0.58 | 9 | **1.09** | **0.024** | 3 | 0.56 | 2 | 0.84 | 0.200 |
| L. Olf. Bulb | 6 | 1.54 | 8 | 2.60 | 0.080 | 3 | 1.07 | 2 | 1.45 | 0.309 |
| L. Subcortical | 10 | 3.10 | 9 | **5.04** | **0.038** | 5 | 2.34 | 4 | **4.11** | **0.034** |
| L. Cortex | 9 | 5.93 | 9 | **11.13** | **0.029** | 4 | 4.58 | 4 | 6.63 | 0.327 |
| Cerebellum | 9 | 1.49 | 9 | 1.74 | 0.491 | 4 | 1.26 | 4 | 1.61 | 0.302 |
| Pons | 10 | 1.77 | 9 | 2.74 | 0.126 | 5 | 1.45 | 4 | 2.01 | 0.254 |
| Medulla | 9 | 5.11 | 9 | 7.13 | 0.109 | 4 | 4.44 | 4 | 7.43 | 0.052 |
| Hypothalamus | 0 | . | 5 | 0.12 | . | 0 | . | 1 | 0.07 | . |
| Choroid Plexus | 10 | 0.22 | 8 | 0.18 | 0.532 | 5 | 0.13 | 4 | 0.41 | 0.327 |
| Supratentorial | 10 | 23.88 | 9 | **47.72** | **0.014** | 5 | 18.08 | 4 | 31.12 | 0.089 |
| Infratentorial | 10 | 7.70 | 9 | 11.68 | 0.072 | 5 | 6.02 | 4 | **11.07** | **0.047** |
| Total Volume | 10 | 32.15 | 9 | **61.23** | **0.014** | 5 | 24.09 | 4 | **43.84** | **0.049** |

** Welch–Satterthwaite t-test was used to test volume differences among individual structures between groups.*

**Supplemental Table 13A: Genotype/Regional Brain Volume: Sap130(m/m), Pcdha9 (*/*) vs WT; Sap130(m/m), Pcdha9 (*/*) vs Sap130(*/+), Pcdha9 (*/+)**

|  | ABC vs. WT | | | ABC vs. F | | |
| --- | --- | --- | --- | --- | --- | --- |
|  | Sap130(m/m), Pcdha9 (*/*) vs WT | | | Sap130(m/m), Pcdha9 (*/*) vs Sap130(*/+), Pcdha9 (*/+) | | |
| Structure | ABC Percent TBV | WT Percent TBV | P value * | ABC Percent TBV | F Percent TBV | P value * |
| R. Hippocampus | 0.015 | 0.018 | 0.238 | 0.015 | 0.017 | 0.328 |
| R. Olf. Bulb | 0.036 | **0.041** | **0.037** | 0.036 | 0.039 | 0.287 |
| R. Subcortical | 0.103 | 0.106 | 0.792 | 0.103 | 0.083 | 0.089 |
| R. Cortex | 0.147 | 0.174 | 0.105 | 0.147 | **0.185** | **0.028** |
| Intraventricular Vol. | 0.049 | 0.011 | 0.063 | 0.049 | 0.034 | 0.420 |
| Midbrain | **0.153** | 0.098 | **0.027** | 0.153 | 0.121 | 0.191 |
| L. Hippocampus | 0.016 | 0.019 | 0.304 | 0.016 | 0.018 | 0.432 |
| L. Olf. Bulb | 0.036 | 0.041 | 0.117 | 0.036 | 0.041 | 0.202 |
| L. Subcortical | 0.106 | 0.107 | 0.898 | 0.106 | 0.083 | 0.086 |
| L. Cortex | 0.157 | 0.171 | 0.430 | 0.157 | 0.183 | 0.138 |
| Cerebellum | 0.043 | 0.043 | 0.864 | **0.043** | 0.030 | **0.010** |
| Pons | 0.056 | 0.048 | 0.102 | 0.056 | 0.045 | 0.104 |
| Medulla | **0.166** | 0.122 | **0.005** | **0.166** | 0.125 | **0.043** |
| Hypothalamus | 0.001 | 0.003 | . | 0.001 | 0.002 | . |
| Choroid Plexus | 0.008 | 0.004 | 0.052 | **0.008** | 0.003 | **0.012** |
| Supratentorial | 0.726 | **1.287** | **0.009** | 0.726 | 0.774 | 0.092 |
| Infratentorial | 0.251 | 0.212 | 0.075 | 0.251 | 0.201 | 0.083 |

** Welch–Satterthwaite t-test was used to test percent volume differences among individual structures between groups.*

**Supplemental Table 13B: Genotype/Regional Brain Volume: Sap130(m/m), Pcdha9 (m/*) vs Sap130(*/+), Pcdha9 (*/+);Sap130(m/m), Pcdha9 (m/m) vs Sap130(m/m), Pcdha9 (+/+)**

|  | AB vs. F | | | A vs. C | | |
| --- | --- | --- | --- | --- | --- | --- |
|  | Sap130(m/m), Pcdha9 (m/*) vs Sap130(*/+), Pcdha9 (*/+) | | | Sap130(m/m), Pcdha9 (m/m) vs Sap130(m/m), Pcdha9 (+/+) | | |
| Structure | AB Percent TBV | F Percent TBV | P value * | A Percent TBV | C Percent TBV | P value * |
| R. Hippocampus | 0.014 | 0.017 | 0.256 | 0.017 | 0.018 | 0.963 |
| R. Olf. Bulb | 0.036 | 0.039 | 0.304 | 0.035 | 0.036 | 0.830 |
| R. Subcortical | 0.105 | 0.083 | 0.168 | 0.119 | 0.097 | 0.486 |
| R. Cortex | 0.148 | 0.185 | 0.132 | 0.150 | 0.147 | 0.940 |
| Intraventricular Vol. | 0.052 | 0.034 | 0.599 | 0.004 | 0.046 | 0.057 |
| Midbrain | 0.164 | 0.121 | 0.195 | 0.195 | 0.127 | 0.324 |
| L. Hippocampus | 0.016 | 0.018 | 0.505 | 0.018 | 0.016 | 0.756 |
| L. Olf. Bulb | 0.038 | 0.041 | 0.529 | 0.032 | 0.028 | 0.344 |
| L. Subcortical | 0.111 | 0.083 | 0.135 | 0.125 | 0.093 | 0.407 |
| L. Cortex | 0.159 | 0.183 | 0.334 | 0.147 | 0.152 | 0.900 |
| Extra-axial CSF | 0.087 | 0.075 | 0.901 | . | 0.050 | . |
| Cerebellum | **0.046** | 0.030 | **0.012** | 0.052 | 0.038 | 0.256 |
| Pons | **0.060** | 0.045 | **0.049** | 0.067 | 0.048 | 0.164 |
| Medulla | 0.165 | 0.125 | 0.091 | 0.184 | 0.169 | 0.679 |
| Hypothalamus | . | 0.002 | . | . | 0.001 | . |
| Choroid Plexus | **0.008** | 0.003 | **0.029** | 0.007 | 0.009 | 0.778 |
| Supratentorial | 0.733 | 0.774 | . | 0.745 | 0.707 | 0.533 |
| Infratentorial | 0.250 | 0.201 | 0.256 | 0.255 | 0.255 | 0.995 |

** Welch–Satterthwaite t-test was used to test percent volume differences among individual structures between groups.*
